# The Utility of Cognitive Testing to Predict Real World Commercial Driving Risk

**DOI:** 10.1101/2022.10.19.22281278

**Authors:** Daniel Scott, Alice Elizabeth Atkin, Aaron Granley, Anthony Singhal

## Abstract

**Background:** Driving is a complex task which requires numerous cognitive and sensorimotor skills to be performed safely. On-road driver evaluation can identify unsafe drivers but can also be expensive, risky, and time-consuming. Poor performance on off-road measures of cognition and sensorimotor control has been shown to predict on-road performance in privately-licensed light vehicle drivers, but commercial drivers have not yet been studied despite such vehicles generally being larger and heavier, thus increasing risks from unsafe driving.

**Method:** Commercially-licensed truck, bus, and light vehicle drivers undertook the tablet-based Vitals cognitive screening tool, which measures reaction time, judgement, memory, and sensorimotor control, and also undertook an on-road driving evaluation using their vehicle. Accuracy and reliability of the Vitals tasks on predicting road test outcomes were investigated using a trichotomous classifier (pass, fail, borderline), and task performance was analyzed depending on vehicle type and road test outcome.

**Results:** Performance on the Vitals tasks predicted on-road performance across all vehicle types. Participants who failed their on-road evaluation also demonstrated lower success on the Judgement task, fewer correctly replicated shapes on the Memory task, and less time on-target in the Control task compared to those who passed.

**Conclusion:** Performance on cognitive and sensorimotor tasks is a good predictor of future driving performance and driver safety for commercially-licensed drivers. Regardless of vehicle type, stakeholders can use cognitive measures from the Vitals assessment to identify an increased driving risk. Use of the Vitals as a screening tool prior to on-road evaluation can benefit both drivers and evaluators.

## 1. Introduction to Cognition, Driving, and Impairment

Driving is an important and complex task which requires various human cognitive and sensorimotor functions in order to maintain safety. For example, safe vehicle operation in traffic requires attention, memory, visual discrimination, object recognition, language, and executive functions such as problem-solving and decision-making (CCMTA, 2022). Impairment to any of these functions may result in reduced performance or an elevated risk of error, resulting in an increased risk of collisions and crashes. Cognitive impairment can have many levels, from mild to severe, and can be either episodic or persistent, depending on the underlying cause. Causes of impairment can include physical injury or disease, psychiatric illness, use of pharmaceutical or recreational drugs such as alcohol or cannabis, as well as situational factors such as active or passive fatigue (Saxby et al., 2013, Hancock & Desmond, 2000). Depending on the nature and severity of impairment, driving performance can sometimes be maintained via compensatory devices or strategies, but for some drivers, the only safe solution to impairment is to cease driving, either temporarily or permanently (CCMTA, 2022).

### 1.1. Driving

Road safety is a significant public health issue all around the world. According to the World Health Organization, as of 2018, traffic fatalities were the leading cause of death in young people aged 5-29 worldwide and the eighth highest cause of death amongst all ages. However, these fatalities are not evenly distributed amongst nations, and traffic fatality rates are up to three times higher in low-income countries compared to high-income countries. In fact, unlike low-income countries, high-income countries generally have experienced a substantial decline in traffic fatality rates; since 2010, every G7 country and the European Union has seen a decline in fatality rates, with the exception of the United States (GHSA, 2022; OECD, 2022). This general decline comes despite progressive aging of the population within high-income countries and a large increase in the number of seniors over the age of 75, who generally match older teenagers (18-20) for the highest rate of traffic fatalities by age group (ITF, 2020; 2021). For instance, the median age of Canadians has increased from 26.2 in 1971 to 41.1 in 2021, and the number of Canadian seniors over the age of 75 has also increased by almost 5-fold (StatsCan, 2021). These national differences in fatality rates and trends suggest that by identifying and addressing relevant factors, the risks of driving can be substantially mitigated, even as the number of at-risk road users increases. Many countries and cities around the world have committed to achieve precisely this via “Vision Zero” pledges which seek to eliminate traffic fatalities and injuries through changes to traffic design and public policy (Johansson, 2009; Kim et al., 2017).

All drivers share the same road infrastructure, but the vehicles that are driven can vary significantly in their design and function. Consequently, driver’s licenses are typically divided into different classes, each requiring separate knowledge and on-road driving tests. There are licenses for operating a personal vehicle, such as an automobile or a motorcycle, and there are also licenses for operating commercial vehicles, such as trucks or buses. Commercial vehicles are typically larger and heavier than personal vehicles, which increases their hazard to other road users (Matsui et al., 2016; Zou et al., 2017). In general, a larger, heavier vehicle will have longer braking distances, larger blindspots, be more difficult to navigate on complex urban streets, and have greater impact energy in the event of a collision. Heavier vehicles tend to be safer for their drivers in the event of a crash, but also increase the risk of injury and death for other road users, especially pedestrians and cyclists (Anderson & Auffhammer, 2014; Li., 2012; Savage, 2013; Schmitt, 2020). Although the large majority of traffic fatalities involve personal vehicles, commercial heavy trucks have a relatively higher fatality rate per vehicle mile, and also incur extremely high costs when crashes occur (FMCSA, 2007; Savage, 2013). In fact, in the United States, commercial truck drivers have some of the highest workplace fatality rates of any occupation (BoLS, 2021), and the nature of the occupation means that commercial truck drivers have notably high rates of health conditions that are associated with an elevated crash risk, such as sleep apnea, hypertension, and diabetes (Abu Dabrh, 2014). Bus crashes are very infrequent and make up only a small percentage of large vehicle crashes in the United States, but their large size and high passenger occupancy make the consequences of a crash especially serious (FMCSA, 2009). Consequently, commercial trucks and buses require a high level of driver skill and vigilance in order to be operated safely, thus necessitating special licensing above and beyond what is required to operate personal vehicles. Furthermore, personal automobiles require skill and vigilance to be operated, making driving performance a good proxy for an individual’s general cognitive health and performance (CCMTA, 2022). A good driver is likely to be in good cognitive health, and a person in good cognitive health is likely to be a good driver.

### 1.2. Driving and Impairment

As a complex task involving numerous cognitive and sensorimotor functions, driving is sensitive to any impairment in these functions, with the level of risk depending upon the level of impairment and its persistence. Every driver is at risk of certain forms of transient impairment, such as fatigue, but for most forms of impairment, the risk tends to be concentrated amongst certain drivers. For instance, government research in Canada and the Netherlands suggests that repeat offenders are responsible for the majority of impaired driving violations and serious crashes involving alcohol (Clermont, 2018; Goldenbeld et al., 2016). Other causes of cognitive impairment, such as neurological, cardiovascular, or metabolic diseases, reliably increase in prevalence with increased age (Dumurgier & Tzourio, 2020; Matthews et al., 2016). For example, in the United States, approximately 0.4% of people under the age of 75 are diagnosed annually with Alzheimer’s disease, but for people over the age of 85, that number is 7.6% (Alzheimer’s Association, 2022). A diagnosis of mild cognitive impairment (MCI), which can be acquired via acute neurological injury but may also be a precursor to dementia, likewise becomes more prevalent with age, as do Type II diabetes (Sharma et al., 2016) and heart disease (Dai et al., 2021). Depending upon the disease and its severity, the resulting impairment can sometimes be minimized via pharmaceuticals and other health interventions, or it can be compensated for via changes in driving behaviour. Some neurological illnesses, such as Alzheimer’s disease, however, are progressive and degenerative, meaning that drivers with these conditions will experience an irreversible deterioration in their cognitive and sensorimotor skills until it is no longer safe to drive. Traditional crash reports suggest that the vast majority of traffic fatalities either involve impairment directly as a factor, such as when drivers are operating their vehicle under the influence of alcohol or drugs, or they can involve driver errors which are substantially more likely to occur if drivers are operating under cognitive impairment (Dingus et al., 2016). Such driver errors come in different forms and are commonly classified as either recognition errors, decision errors, or performance errors (Singh, 2015). A recognition error is when a driver fails to perceive or attend to some critical aspect of the road environment, such as a stop sign, traffic light, or pedestrian. A decision error is when a driver makes a decision that deviates from safe or legal driving practices, such as speeding, tailgating, or making unsafe maneuvers across traffic lanes. A performance error is when a driver fails to properly execute an intended driving maneuver, such as reacting slowly to an obstacle or turning the vehicle imprecisely. A narrow focus on driver error as the cause of most crashes has been criticized by the Vision Zero approach to traffic safety, which instead adopts a “systems perspective” (Kim et al., 2017) to crash analysis by assessing driver behaviour in the wider context of vehicle design, road design, and transportation policy, and by advocating for vehicles and roads which are more tolerant of error (Johansson, 2009; Schmitt, 2020). Nevertheless, it remains the case that driver errors are more likely to occur when a driver is cognitively impaired, particularly by alcohol (Compton & Berning, 2015; Dingus et al., 2016) and cannabis (Bondallaz et al., 2016; Compton, 2017; Hartman & Huestis, 2013), and that some policies to reduce impaired driving have proven successful and are globally recommended, such as lowering legal blood alcohol concentration (BAC) limits, as well as enforcement and points systems which can lead to license suspensions or criminal penalties for drivers who exceed those limits (Castillo-Manzano & Castro-Nuño, 2012; Fell & Scherer, 2017; WHO, 2021).

### 1.3. License Renewal and Driver Screening

Concerns about impaired driving mean that in addition to administering tests when drivers first acquire their license, many jurisdictions now require certain drivers to have their on-road driving skills regularly re-tested as a condition of license renewal. For instance, in the province of Alberta, Canada, drivers must pass a road test at age 75, age 80, and then every two years thereafter (Government of Alberta, 2022a). This age-based testing schedule is even stricter for commercial drivers, with such tests taking place every five years until age 45, every two years until age 65, and then every year thereafter. Road tests are also often required for drivers who have their licenses suspended after a drug or alcohol-related traffic offense (Government of Alberta, 2022b), and medical professionals can be empowered to recommend that a patient undertake a road test whenever impairment is suspected due to poor health (Government of Alberta, 2022c). While license renewal programs for older adults are intended to identify cognitively impaired drivers and remove them from the road, the evidence of their effectiveness is often found to be minimal (Ichikawa et al., 2015; Langford et al., 2004; Mitchell, 2008; Rock, 1998; Vanlaar et al., 2016). Since it is undoubtedly the case that drivers who are at a high risk of crashing due to impairment or frailty should not be driving, the questionable effectiveness of currently extant license renewal programs and the broad burden that they impose on drivers and jurisdictions suggests a potential role for off-road assessments of cognitive and sensorimotor skills which are important for driving, and which can reliably recommend drivers for an additional on-road test based on individual rather than population-level risk.

One established method of driver screening is the DriveABLE Cognitive Assessment Tool (DCAT), created by Impirica. The DCAT, first introduced in 1997, includes a variety of tasks that measure cognitive ability and can be administered via computer in less than 30 minutes (Dobbs, 1997). Its revised version, Vitals, can be administered on a tablet in the same amount of time. These tasks include measures of reaction time, decision-making, working memory, and sensorimotor coordination and are conceptually related to the forms of driver error (recognition, decision, and performance errors) commonly implicated in crashes (CCMTA, 2022; Impirica, 2022). The DCAT is employed as an assessment tool in Canada by a number of healthcare systems and transportation companies, and performance on the DCAT and Vitals has shown good predictive value in several studies for whether drivers will subsequently pass or fail a road test (Dobbs, 1997; 2013; Dobbs et al., 1998; Korner-Bitensky & Sofer, 2009), particularly within groups of patients who have been diagnosed with a specific neurological condition such as dementia (Bakhtiari et al., 2020) or stroke (Choi et al., 2015).

Two recent studies from our research group will serve to illustrate the potential utility of Vitals delivered on a handheld tablet as a screening test for at-risk drivers. This test battery is comprised of tasks to test for simple reaction time, attention shifting, decision-making, memory, and sensorimotor control. In one study, we demonstrated that drivers who were identified by their physician as being at-risk for unsafe driving performed worse on the test battery relative to healthy controls (Bakhtiari et al., 2020). Furthermore, poorer performance on the test battery in healthy older adults strongly predicted subsequent failure on an on-road driving test. In a second study, we have found that Vitals performance differed in adults under the influence of cocaine, cannabis, or both cannabis and cocaine (Tomczak et al., n.d.). The results showed differential sensitivity to various cognitive factors depending on which drug group they were in, and in comparison to adults not under the influence of drugs or alcohol. We made the key assumption that none of the drug influenced individuals were fit to drive. Together, these two studies indicate that drivers who are likely to pose a considerable risk to themselves or others, and who are likely to fail an on-road driving test, can be reliably identified using an off-road, tablet-based screening tool that can be administered relatively quickly, efficiently, and at low cost. We believe a reliable and widely used computer or tablet-based screening tool could significantly reduce the regulatory burden on drivers and improve the effectiveness of license renewal programs by dissociating cognitively healthy and at-risk individuals prior to a road test, with road tests reserved only for drivers at high risk of unsafe driving and test failure.

### 1.4. Current Study

In this study, we had two main purposes. First, we sought to examine performance differences on the Vitals tablet-based cognitive test battery for commercial vehicle licensing by administering it to commercially licensed truck, bus, and automobile drivers. As described in the foregoing review, these commercial vehicle types differ in many ways, and the driver demands of the different vehicles are unique. It remains an open question whether trained drivers in each of these categories have a different relationship between cognitive performance and on-road driving success. The second purpose of this study was to validate the overall predictive value of the Vitals tablet-based cognitive test battery for on-road performance across all commercial driving categories. The strength of this approach is the large number of drivers that can be used for multivariate predictability. Truck and bus drivers pose a particular safety risk when cognitive impairment is suspected, given the large size of their vehicles and the higher level of skill required to operate them. The stricter testing schedule that commercial drivers face when seeking to retain their license also means that the benefits of a computer-based screening test may be especially high for these drivers, relative to drivers who are only licensed to operate personal vehicles. Thus, in this study, we had commercial truck, bus, and small vehicle drivers perform the Vitals cognitive test battery and also perform a vehicle-specific on-road test. We examined their performance on each dependent measure of the cognitive tests and used the aggregate data to explore the predictability of cognitive task performance to overall commercial driving success.

## 2. Methods

### 2.1. Participants

We compared data from three separately recruited populations associated with different commercial vehicle types. Table 2 contains the available demographic data for each of the groups. The first group was Heavy vehicles, consisting of 415 adult participants (mean age = 41.7 yrs., SD = 13.0). The second group was the Light vehicles, including car, van, and light commercial truck drivers consisting of 1592 adult participants (mean age = 36.95 yrs., SD = 10.7). The third group was Trucking, consisting of 160 adult participants (mean age = 43.69 yrs., SD = 12.57). Due to procedural constraints, we were unable to accurately record the gender of the participants. However, we are confident that most of the participants were male in all three groups. The median time spent on the on-road driving assessment was 64 minutes, although the time distributions varied across organizations. Differences in time were best explained by the unique road constraints required for different vehicle types. For example, large trucks were limited to driving on designated trucking roads, but small vehicles were not.

This study was conducted according to the Declaration of Helsinki (1996). It was approved by the University of Alberta Health Research Ethics Board and performed in compliance with relevant laws and institutional guidelines. All participants gave informed consent to have their data collected for this study. Completion of the Vitals assessment and on-road assessment was linked to each participant’s job duties or was performed as a condition of their hiring by an organization, but participants had the option of completing the evaluations whilst refusing consent to have their data included in this study. The study took approximately 90 minutes to complete – 30 minutes for the Vitals and 60 minutes for the on-road assessment.

### 2.2. Procedure

The Vitals cognitive screen was based on decades of cognitive science research and designed to test the primary cognitive, perceptual, and action-related measures intimately related to driving performance (Bakhtiari et al., 2020). Its tasks include reaction time (RT), decision-making (Judgement), Memory, and sensorimotor control (Control). The Commercial On-Road Evaluation (CORE) was designed to test driving ability judged by a trained instructor for each of the vehicle types. Figure 1 shows the process employed for data collection for both the Vitals and CORE assessments.

**Figure 1:**
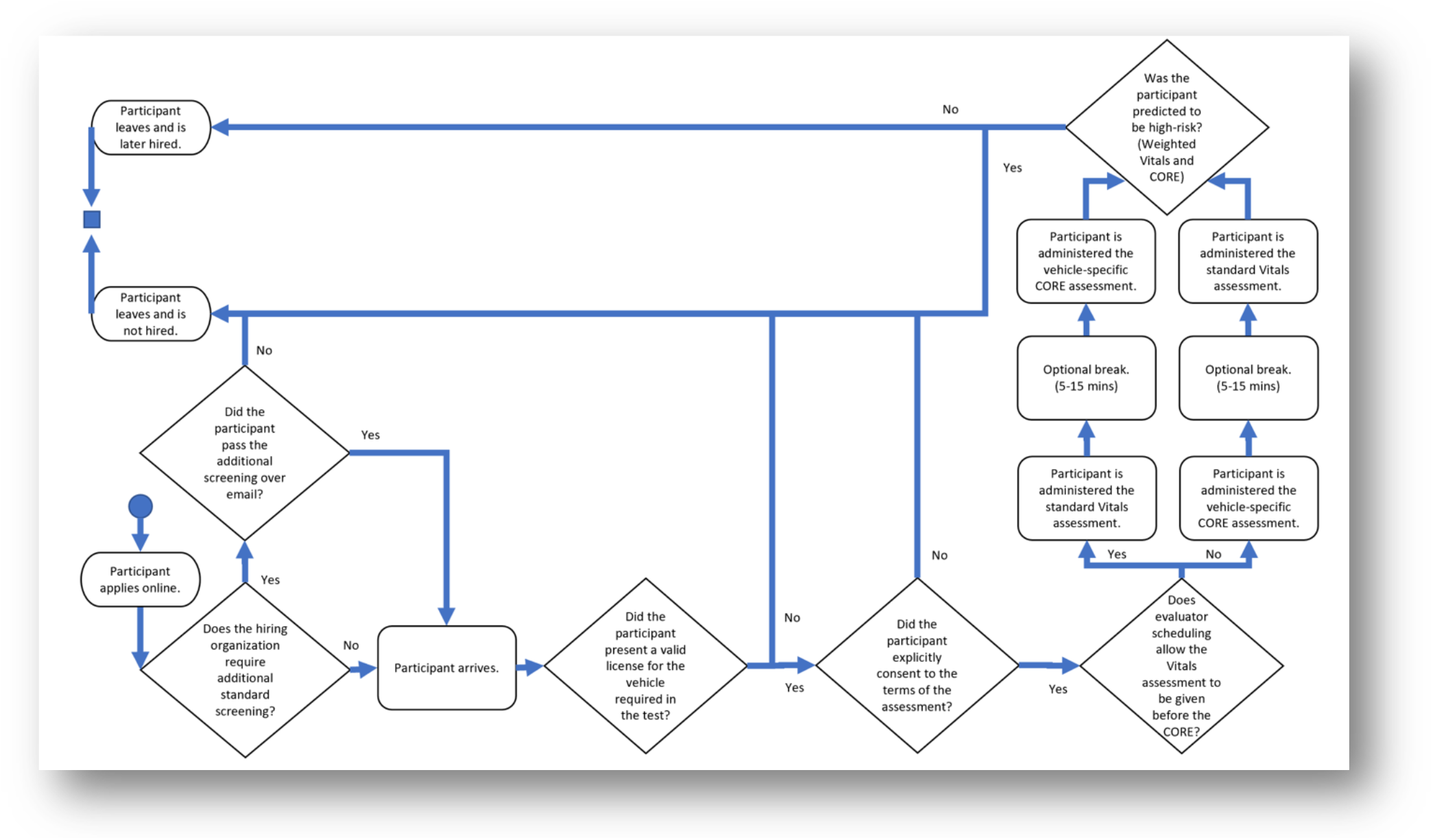
Unified Modelling Language (UML) diagram with detail of the participant screening process and decision tree for the CORE/Vitals pre-hiring procedure. Participants who were being assessed as part of a performance evaluation, for periodic wellness, or after an at-work incident, may have been subject to additional screening related to the continuation of their duties.

### 2.3. Tasks

#### 2.3.1. Cognitive Tasks (Vitals)

The four manually controlled, tablet-based tasks included in this study were designed and developed by Impirica to run as a continuous ordered series. These four tasks included:

1. Reaction Time (RT): participants press a button as fast as possible after a visual cue. The cue can be spatially congruent with the button location or incongruent. Thus, this task also tests for attention switching.
2. Judgement, i.e., decision-making: participants press a ‘Go’ button after a visual cue while avoiding moving obstacles. A ‘Stop’ button is given as an additional control to avoid obstacles. There are two stages. Stage one has one set of obstacles and the second stage has two sets of obstacles. There are ten trials in the first stage and 20 trials in the second stage.
3. Memory: participants draw a previously presented geometric shape with their finger. There are four stages, with four trials in each stage. The number of shapes to recall, shape complexity, and the duration of an intervening mask increases as the stages progress.
4. Control, i.e., bi-manual sensorimotor control: participants follow a target circle using the iPad as if it was a steering wheel to avoid fixed and unpredictable moving obstacles. There are four stages, where speed increases as the stages progress.

Vitals uses reliable measures of cognitive processes known to be associated with safe driving and are predictive of on-road performance in cognitively impaired drivers (Dobbs, 1997; 2013; Dobbs et al., 1998; Bakhtiari et al., 2020). The RT task is a modified version of an attentional shifting task (Monsell, 2003; Robbins, 2007), while the Judgement task is related to other tasks of spatial judgement. The Memory task is a modified version of the Corsi block test (Kessels et al., 2000). The Control task is chosen because it requires similar sensorimotor coordination to that of on-road driving.

In designing Vitals, tests were selected to cover most of the required cognitive domains needed for driving (CCMTA, 2022; Dobbs, 1997); divided attention (the ability to attend to two or more stimuli at the same time, evaluated in bi-manual sensorimotor task), selected attention (the ability to selectively attend to one or more important stimuli while ignoring competing stimuli, evaluated in reaction speed and decision-making tasks), sustained attention (the capacity to maintain attentional activity over a period of time, evaluated by bi-manual sensorimotor task), short-term memory (the temporary storage of information that is currently being processed in a person’s mind, evaluated by memory task), working memory (the ability to manipulate information with time constraints/taking in and updating information to solve problems, evaluated by all four tasks), complex reaction time (the time taken to respond differentially to two or more stimuli or events, evaluated by reaction speed and decision-making tasks), tracking (the ability to visually follow a stimulus that is moving or sequentially appearing in different locations, evaluated by bi-manual sensorimotor task), visuospatial abilities (processes dependent on vision such as the recognition of objects, the ability to mentally rotate objects and determination of relationships between stimuli based on size or color, evaluated by decision-making task), executive functioning (capabilities that enable an individual to successfully engage in independent, purposeful, and self-serving behaviors, evaluated by all four tasks), and visual information processing (the processing of visual information beyond the perceptual level, evaluated by all four tasks).

The Vitals cognitive test battery is administered by an evaluator that is trained and certified. Each task includes a demonstration and practice. If the administrator evaluates that an individual does not comprehend a task during its demonstration, the demonstration is repeated to aid comprehension but not to improve performance. If the administrator evaluates that the participant comprehends the task, but performs poorly, the demonstration is not repeated. After the demonstration is complete, the administrator can answer the participant’s questions with the provided standard responses from a manual.

#### 2.3.2. Commercial On-Road Evaluation (CORE)

The CORE is a standardized on-road driving assessment with a trained driving instructor. The evaluation takes approximately 60 minutes to complete and is based on a scientifically developed on-road evaluation designed to test for a decline in the cognitive skills necessary for safe driving (Berndt et al., 2007; Dobbs et al., 1998; Kowalski & Tuokko, 2007). The CORE is a modification of the DriveABLE On-Road Evaluation (DORE), which was previously reported in Bakhtiari et al. (2020). The CORE assesses the same primary error categories as the DORE but includes additional categories depending on each participant’s vehicle type. This is due to physical differences in vehicle size and controls, including brakes, maneuverability, blind spots, and necessary visual scanning. The total difference in errors between vehicle types is 4% (three differences across 70 error categories).

### 2.4. Vitals Dependent Measures

For each participant, dependent measures were extracted from the Vitals cognitive test battery. Table 1 contains a list of dependent measures from each task. They are as follows. From the RT task, the average reaction times (Reaction time) were extracted. From the Judgement task, the percentage of trials that participants started early (% Premature start), the percentage of trials successfully passing the moving obstacles (% Success), the number of times pressing the ‘Start’ button (Start count), the average time taken to press “Go” after the visual cue (Reaction time), and the average number of obstacles that passed after the visual cue and before finishing passing the obstacles (Obstacle count). From the Memory task, duration of task completion (Duration) and the percentage of correct shape retrieval (% Correct shape) were extracted. From the Control task, the percentage of time inside the target (% Time inside target) and the percentage of times that surprise obstacles were avoided (% Surprise obstacles avoided) were extracted. All dependent measures have their non-linear effects of age removed prior to analysis and integration into models. Age effects were explained and removed using a polynomial function fit to 70% of the training dataset (940 of 1343 training samples) and tested for stability on the remaining 30% (403 of 1343 training samples) used for model testing within the training dataset. Similarly, the analysis performed on the validation dataset removes age effects but only uses the function estimate’s fit on the training dataset.

**Table 1:** Tasks and dependent measures from Vitals.

| Task | Measure | Description |
| --- | --- | --- |
| Reaction Time | Reaction time | Average reaction time to press “Stop” button after the visual cue |
| Judgement | % Premature starts | Percentage of releasing “Start” button before the visual cue |
|  | % Success | Percentage of successfully navigating through the moving obstacles |
|  | Start count | Average number of times “Start” button pressed |
|  | Reaction time | Average time taken to press “Go” after the visual cue |
|  | Obstacle count | Average number of passed obstacles, prior to engagement |
| Memory | Duration | Average time to complete task |
|  | % Correct shape | Percentage of shapes drawn correctly |
| Control | % Time inside target | Percentage of time inside the target |
|  | % Surprise obstacles avoided | Percentage of surprise obstacles avoided |

**Table 2:** On-road pass/borderline/fail sample sizes for drivers of heavy vehicles, light vehicles, and trucks in the validation dataset (n=2167).

|  | Driving Pass | Driving Borderline | Driving Fail | N | Age (M ± SD) |
| --- | --- | --- | --- | --- | --- |
| Heavy | 273 | 65 | 77 | 415 | 41.7 ± 13.0 |
| Light | 919 | 346 | 327 | 1592 | 36.95 ± 10.7 |
| Trucking | 99 | 22 | 39 | 160 | 43.69 ± 12.57 |

### 2.5. Statistical Analyses

To validate the Vitals assessment’s power to predict the CORE pass/fail outcome, we evaluated the performance of our logistic regression model across the three distinctly identifiable vehicle categories (Light, Heavy, and Trucking). All the model’s parameters were fit on a training dataset prior to the validation dataset collection. A logistic model’s decision cutpoint can be calibrated to balance the diagnostic tradeoffs of misclassifying positive and negative outcomes. When there are asymmetric consequences between correct and incorrect predictions, two cutpoints (trichotomous) can be used instead of one (dichotomous) to limit decision sensitivity to within a certain threshold of confidence. If a predicted probability of driving risk is between an upper and lower trichotomous threshold, it is set aside as an undefined decision. This protects all stakeholders from downside when only under-confident predictions are available from the model. The Vitals assessment was designed to address real-world driving risks and must minimize the potential hazard shared by the individual and the traffic system (Bakhtiari et al., 2020). To address this, we applied a set of trichotomous thresholds which are fit to maximize both positive predictive value (likelihood of on-road failure if predicted to be high-risk on Vitals), and negative predictive value (likelihood of on-road pass if predicted to be low-risk on Vitals) measures prior to the collection of the validation dataset. The cutpoints were optimized on the training dataset (n=1343) using the pseudo minimization objective (FPR x “false positive penalty”) + (FNR x “false negative penalty”). To define our cutpoint objective, distinct cases of misclassification were first identified by industry experts (e.g. participant received very high Vitals score but very good on-road performance) in the training dataset and assigned a penalty. The penalties quantified negative consequences across dimensions of participant mental hazard (false positive) and driving accident cost (false negative). Since the objective’s penalties are human labeled, discrete, and non-differentiable, a direct search optimization method is advised (Silva et al., 2018). The Nelder-Mead simplex method was selected because of its usefulness for non-differentiable unconstrained objective optimization (Silva et al., 2018). Alternatives were evaluated as the sum of distinct misclassifications multiplied by the rate of their respective occurrence across the training dataset for a given set of cutpoints. Alternatives that resulted in undefined decision rates larger than 20% were discarded. The final trichotomous cutpoint solution which minimized the objective was 0.48 and 0.68 for the low and high cutpoints, respectively. This set of trichotomous thresholds was used for the evaluation of the validation dataset (n=2167) and was not refit once the validation data collection began.

Demographic data of the sample used to generate the classifier model are shown in Table 9. A variety of data was used, including commercial, medical, judicial, and academic sources.

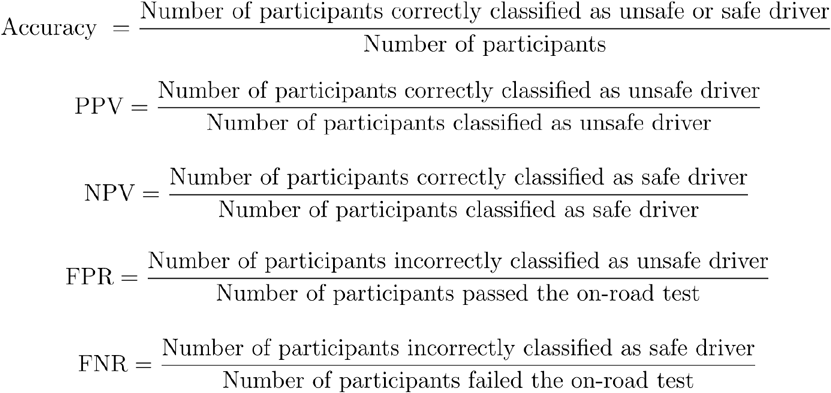

## 3. Results

### 3.1. Descriptive Statistics

Table 2 presents on-road test outcomes for each driving group (Light, Heavy, and Trucking). Based on results from the standard on-road evaluation test, drivers who drove safely or dangerously were grouped as pass or fail, respectively. Participants whose driving errors were toward the upper end, but did not exceed, the range for safe drivers are categorized as borderline. Table 3 presents the mean, standard deviation, and median scores on each measure of the Vitals tasks for the three driving groups. Figure 2 presents data regarding the purpose of assessment for participants within the three driving groups. The majority of participants in each driving group were recruited to the study as part of a pre-hire evaluation for prospective employees, however, the Heavy vehicle group did include a disproportionate number of participants who were assessed as part of a performance evaluation or as part of a return-to-work process following a workplace incident. Individuals that were unable to complete a task for any reason were excluded from the task’s analysis.

**Table 3:**
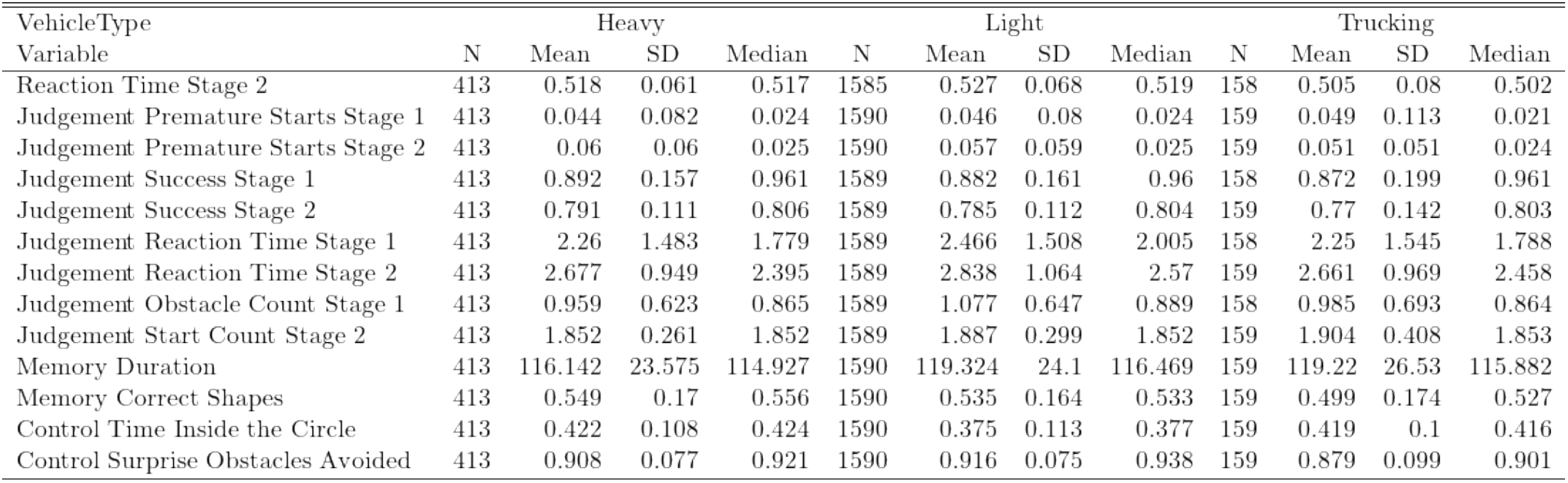
Mean and median performance on Vitals tasks by driving group (Heavy, Light, and Trucking) in the validation dataset (n=2167).

| VehicleType | Heavy |  |  |  | Light |  |  |  | Trucking |  |  |  |
| --- | --- | --- | --- | --- | --- | --- | --- | --- | --- | --- | --- | --- |
| Variable | N | Mean | SD | Median | N | Mean | SD | Median | N | Mean | SD | Median |
| Reaction Time Stage 2 | 413 | 0.518 | 0.061 | 0.517 | 1585 | 0.527 | 0.068 | 0.519 | 158 | 0.505 | 0.08 | 0.502 |
| Judgement Premature Starts Stage 1 | 413 | 0.044 | 0.082 | 0.024 | 1590 | 0.046 | 0.08 | 0.024 | 159 | 0.049 | 0.113 | 0.021 |
| Judgement Premature Starts Stage 2 | 413 | 0.06 | 0.06 | 0.025 | 1590 | 0.057 | 0.059 | 0.025 | 159 | 0.051 | 0.051 | 0.024 |
| Judgement Success Stage 1 | 413 | 0.892 | 0.157 | 0.961 | 1589 | 0.882 | 0.161 | 0.96 | 158 | 0.872 | 0.199 | 0.961 |
| Judgement Success Stage 2 | 413 | 0.791 | 0.111 | 0.806 | 1589 | 0.785 | 0.112 | 0.804 | 159 | 0.77 | 0.142 | 0.803 |
| Judgement Reaction Time Stage 1 | 413 | 2.26 | 1.483 | 1.779 | 1589 | 2.466 | 1.508 | 2.005 | 158 | 2.25 | 1.545 | 1.788 |
| Judgement Reaction Time Stage 2 | 413 | 2.677 | 0.949 | 2.395 | 1589 | 2.838 | 1.064 | 2.57 | 159 | 2.661 | 0.969 | 2.458 |
| Judgement Obstacle Count Stage 1 | 413 | 0.959 | 0.623 | 0.865 | 1589 | 1.077 | 0.647 | 0.889 | 158 | 0.985 | 0.693 | 0.864 |
| Judgement Start Count Stage 2 | 413 | 1.852 | 0.261 | 1.852 | 1589 | 1.887 | 0.299 | 1.852 | 159 | 1.904 | 0.408 | 1.853 |
| Memory Duration | 413 | 116.142 | 23.575 | 114.927 | 1590 | 119.324 | 24.1 | 116.469 | 159 | 119.22 | 26.53 | 115.882 |
| Memory Correct Shapes | 413 | 0.549 | 0.17 | 0.556 | 1590 | 0.535 | 0.164 | 0.533 | 159 | 0.499 | 0.174 | 0.527 |
| Control Time Inside the Circle | 413 | 0.422 | 0.108 | 0.424 | 1590 | 0.375 | 0.113 | 0.377 | 159 | 0.419 | 0.1 | 0.416 |
| Control Surprise Obstacles Avoided | 413 | 0.908 | 0.077 | 0.921 | 1590 | 0.916 | 0.075 | 0.938 | 159 | 0.879 | 0.099 | 0.901 |

**Figure 2:**
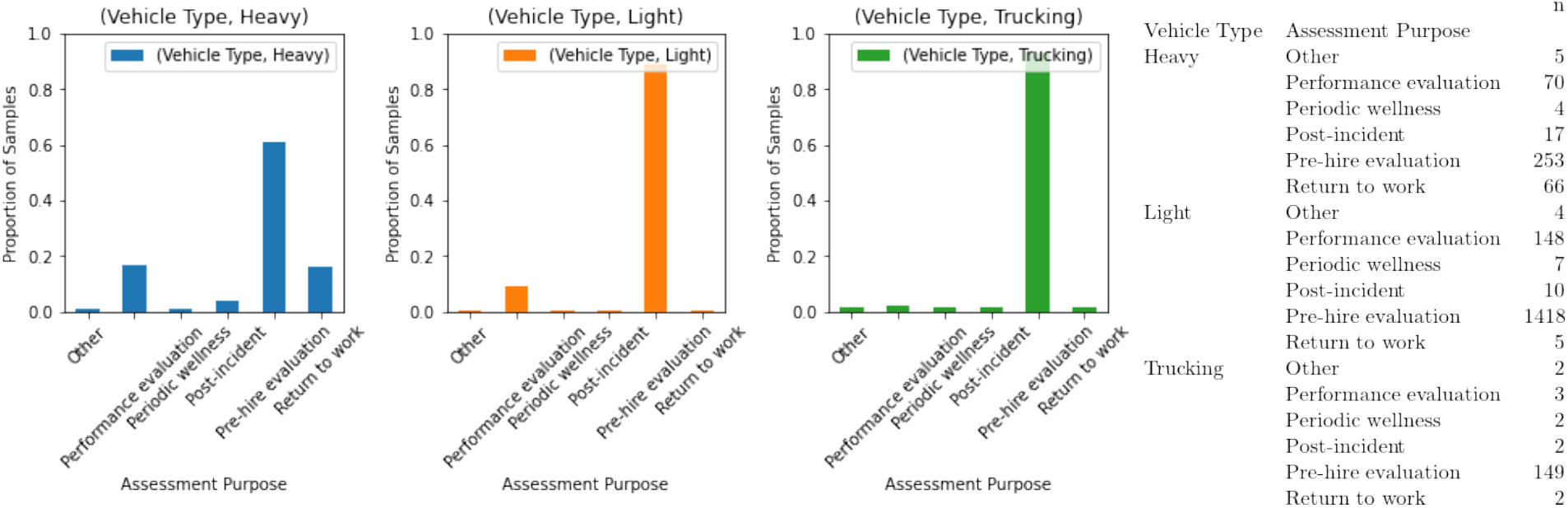
Purpose of assessment for participants within each driving group in the validation dataset (n=2167). Figures 2a, 2b, and 2c display each recorded assessment purpose as a proportion of assessments for each driving group. Figure 2d displays each recorded assessment purpose as a raw count for all driving groups.

### 3.2 Group Comparisons

#### 3.2.1 Full Sample

As seen in Table 4 and Table 5, participants were divided into pass and fail groups. Performance on each measure of the Vitals tasks was compared between groups. Participants who failed their on-road evaluation were significantly less successful on Stage 2 of the Judgement task, performed fewer false starts on Stage 2 of the Judgement task, replicated fewer shapes on the Memory task, and spent less time within the target circle on the Control task.

**Table 4:** Bonferroni-adjusted t-test comparison of means by on-road test outcome (fail vs. pass) in the validation dataset (n=2167).

|  | Variables | fail (n) | pass (n) | statistic | df | p | p.adj | sig |
| --- | --- | --- | --- | --- | --- | --- | --- | --- |
| 1 | Reaction Time Stage 2 | 443 | 1724 | 1.41 | 601.88 | 0.16 | 1.00 | ns |
| 2 | Judgement Premature Starts Stage 1 | 443 | 1724 | 1.94 | 611.00 | 0.05 | 0.68 | ns |
| 3 | Judgement Premature Starts Stage 2 | 443 | 1724 | 1.61 | 575.22 | 0.11 | 1.00 | ns |
| 4 | Judgement Success Stage 1 | 443 | 1724 | -2.44 | 605.78 | 0.01 | 0.20 | ns |
| 5 | Judgement Success Stage 2 | 443 | 1724 | -6.09 | 530.93 | 0.00 | 0.00 | **** |
| 6 | Judgement Reaction Time Stage 1 | 443 | 1724 | 0.30 | 668.45 | 0.76 | 1.00 | ns |
| 7 | Judgement Reaction Time Stage 2 | 443 | 1724 | 2.25 | 642.37 | 0.02 | 0.32 | ns |
| 8 | Judgement Obstacle Count Stage 1 | 443 | 1724 | 0.26 | 663.46 | 0.79 | 1.00 | ns |
| 9 | Judgement Start Count Stage 2 | 443 | 1724 | -3.22 | 698.74 | 0.00 | 0.02 | * |
| 10 | Memory Duration | 443 | 1724 | 2.83 | 660.61 | 0.00 | 0.06 | ns |
| 11 | Memory Correct Shapes | 443 | 1724 | -4.30 | 647.47 | 0.00 | 0.00 | *** |
| 12 | Control Time Inside the Circle | 443 | 1724 | -6.02 | 713.84 | 0.00 | 0.00 | **** |
| 13 | Control Surprise Obstacles Avoided | 443 | 1724 | -0.55 | 694.12 | 0.58 | 1.00 | ns |

**Table 5:** Wilcoxon comparison of medians by on-road test outcome (fail vs pass), using Holm adjustment in the validation dataset (n=2167).

|  | Variables | fail (n) | pass (n) | statistic | p | p.adj | sig |
| --- | --- | --- | --- | --- | --- | --- | --- |
| 1 | Reaction Time Stage 2 | 443 | 1724 | 399745.00 | 0.06 | 0.39 | ns |
| 2 | Judgement Premature Starts Stage 1 | 443 | 1724 | 391462.00 | 0.33 | 1.00 | ns |
| 3 | Judgement Premature Starts Stage 2 | 443 | 1724 | 394198.00 | 0.23 | 1.00 | ns |
| 4 | Judgement Success Stage 1 | 443 | 1724 | 363206.00 | 0.17 | 1.00 | ns |
| 5 | Judgement Success Stage 2 | 443 | 1724 | 319058.50 | 0.00 | 0.00 | **** |
| 6 | Judgement Reaction Time Stage 1 | 443 | 1724 | 381691.00 | 0.82 | 1.00 | ns |
| 7 | Judgement Reaction Time Stage 2 | 443 | 1724 | 405450.00 | 0.03 | 0.20 | ns |
| 8 | Judgement Obstacle Count Stage 1 | 443 | 1724 | 378836.00 | 0.99 | 1.00 | ns |
| 9 | Judgement Start Count Stage 2 | 443 | 1724 | 345039.00 | 0.00 | 0.03 | * |
| 10 | Memory Duration | 443 | 1724 | 409045.00 | 0.01 | 0.12 | ns |
| 11 | Memory Correct Shapes | 443 | 1724 | 333888.50 | 0.00 | 0.00 | *** |
| 12 | Control Time Inside the Circle | 443 | 1724 | 312453.00 | 0.00 | 0.00 | **** |
| 13 | Control Surprise Obstacles Avoided | 443 | 1724 | 370326.00 | 0.40 | 1.00 | ns |

On each measure of the Vitals tasks, participant performance was compared depending on their driving group and whether they passed or failed the on-road evaluation. For the RT task (Figure 3), light vehicle drivers who passed the test had a significantly faster average RT than light vehicle drivers who failed. No differences were observed for truck or heavy vehicle drivers depending on if they passed or failed their test.

**Figure 3:**
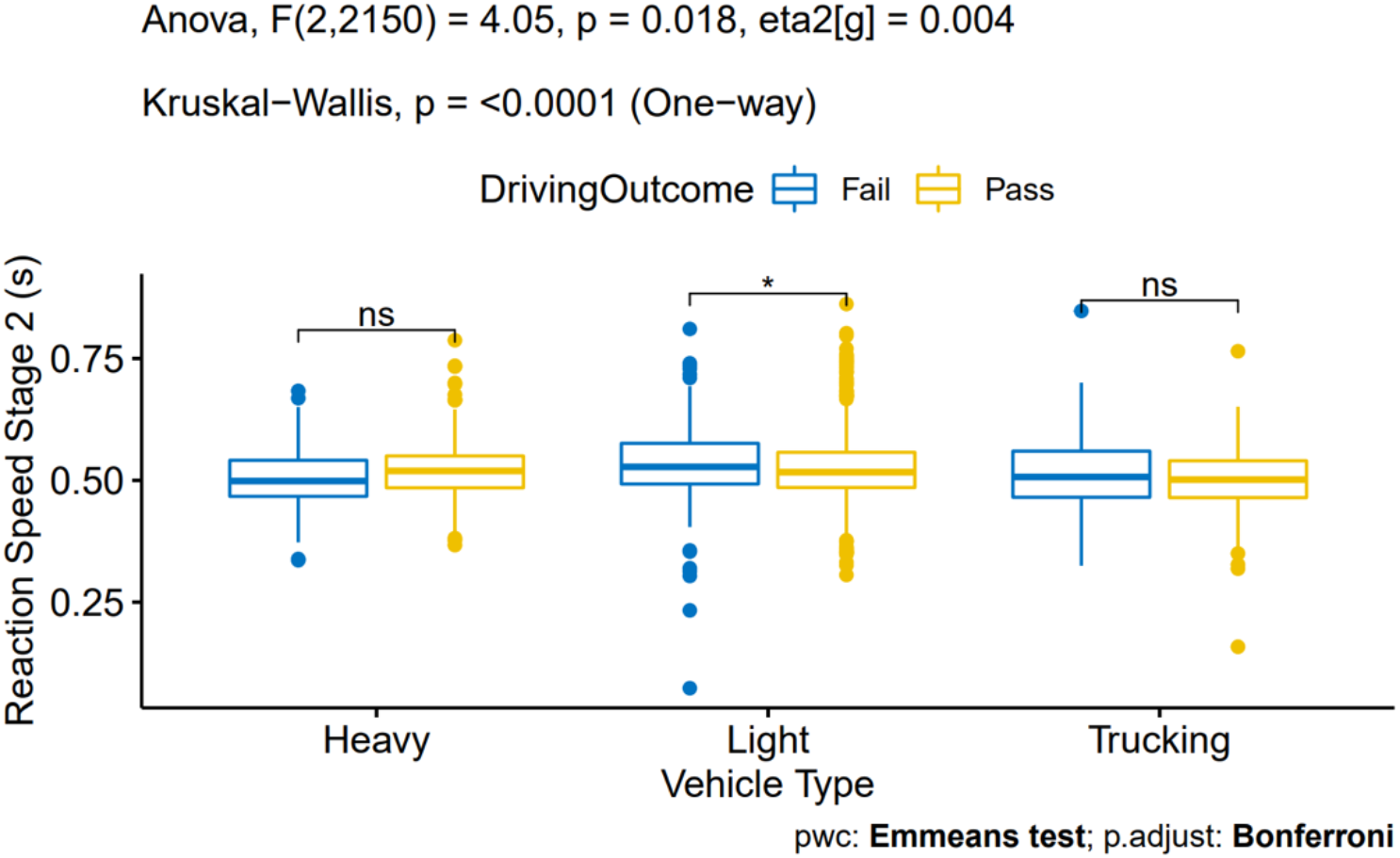
Group means and differences comparing reaction time on the RT task depending on driving group and on-road test outcome. Light vehicle drivers who failed the road test were slower than light vehicle drivers who passed.

On Stage 1 of the Judgement task, light vehicle drivers who passed the on-road test made significantly fewer premature starts (Figure 4) and were more successful (Figure 5) than drivers who failed. No differences were observed for truck or heavy vehicle drivers depending on if they passed or failed their test. No differences in average RT or the number of obstacles passed prior to engagement were observed for any driving group, depending on if they passed or failed. On Stage 2 of the task, light vehicle drivers who passed the on-road test were significantly more successful (Figure 6) than light vehicle drivers who failed and made more starts and stops while avoiding obstacles (Figure 8). Truck drivers who passed the on-road test were also significantly more successful (Figure 6) and had a faster average RT (Figure 7) than truck drivers who failed. No differences were observed for heavy vehicle drivers on any measure depending on if they passed or failed their test. No differences were observed in the number of premature starts for any driving group.

**Figure 4:**
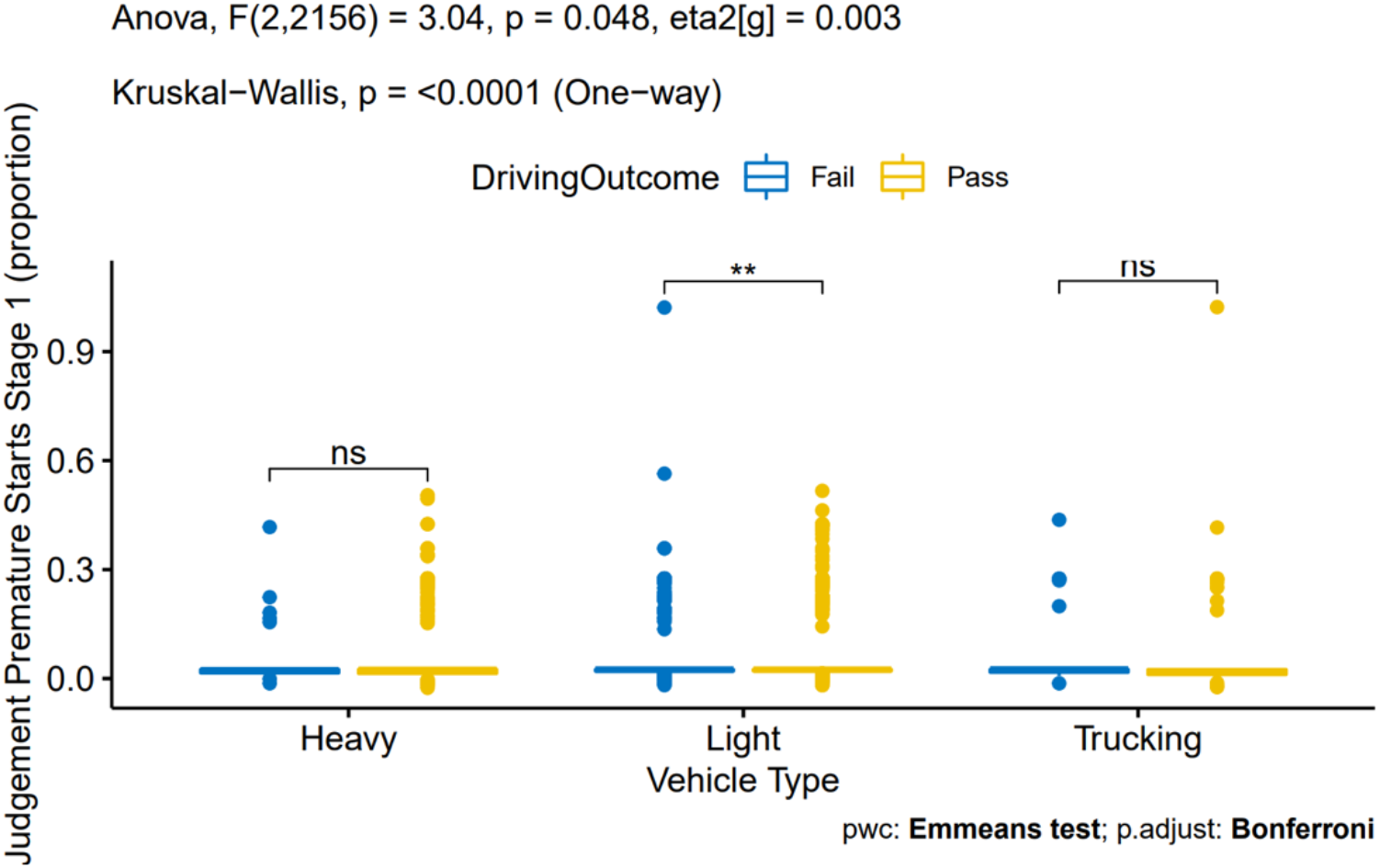
Group means and differences comparing proportion of total starts that were premature on Stage 1 of the Judgement task, depending on driving group and on-road test outcome. Light vehicle drivers who passed the test produced fewer false starts than light vehicle drivers who failed as a proportion of starts.

**Figure 5:**
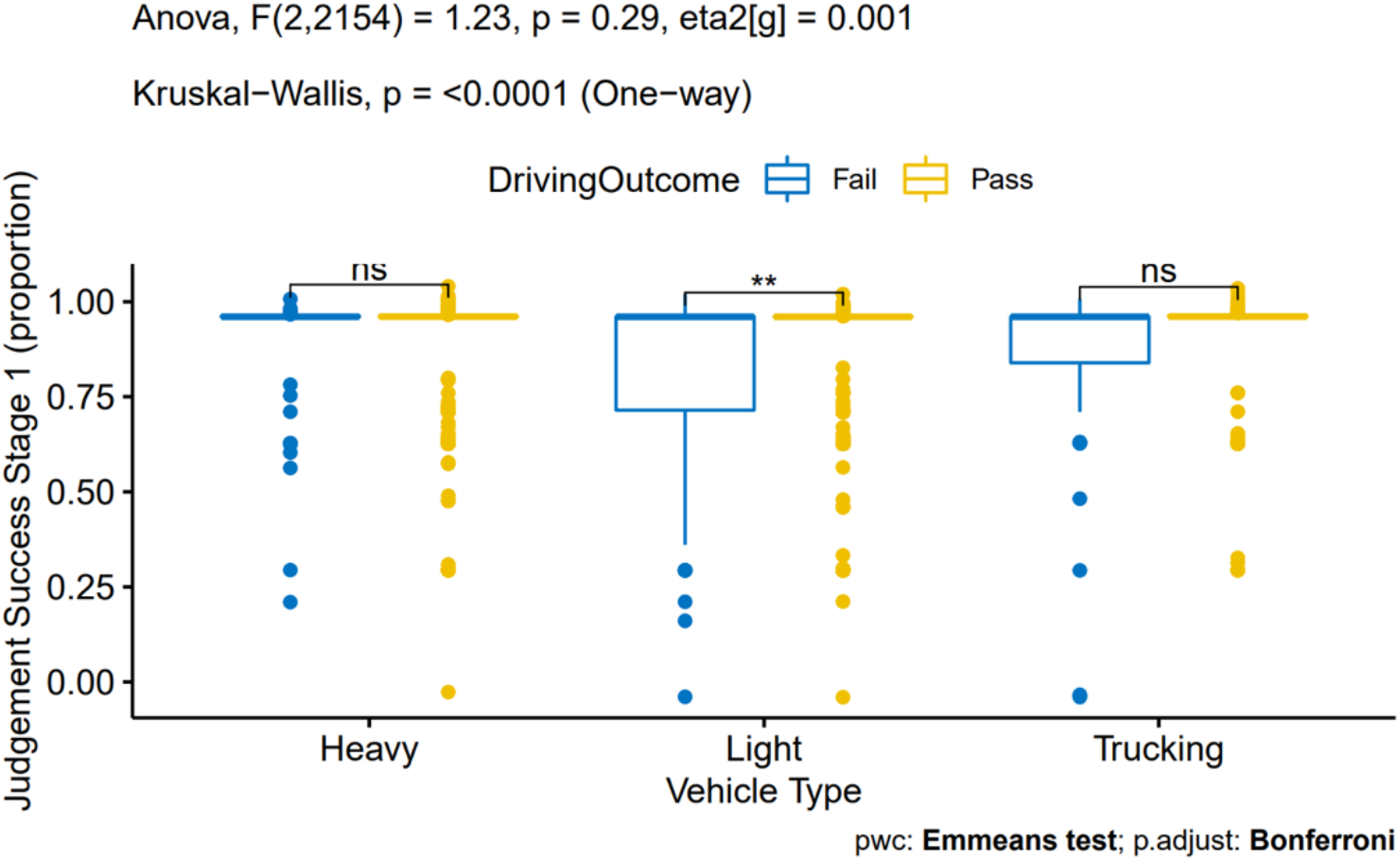
Group means and differences comparing trial success rate on Stage 1 of the Judgement task, depending on driving group and on-road test outcome. Light vehicle drivers who passed the on-road test were more successful than light vehicle drivers who failed.

**Figure 6:**
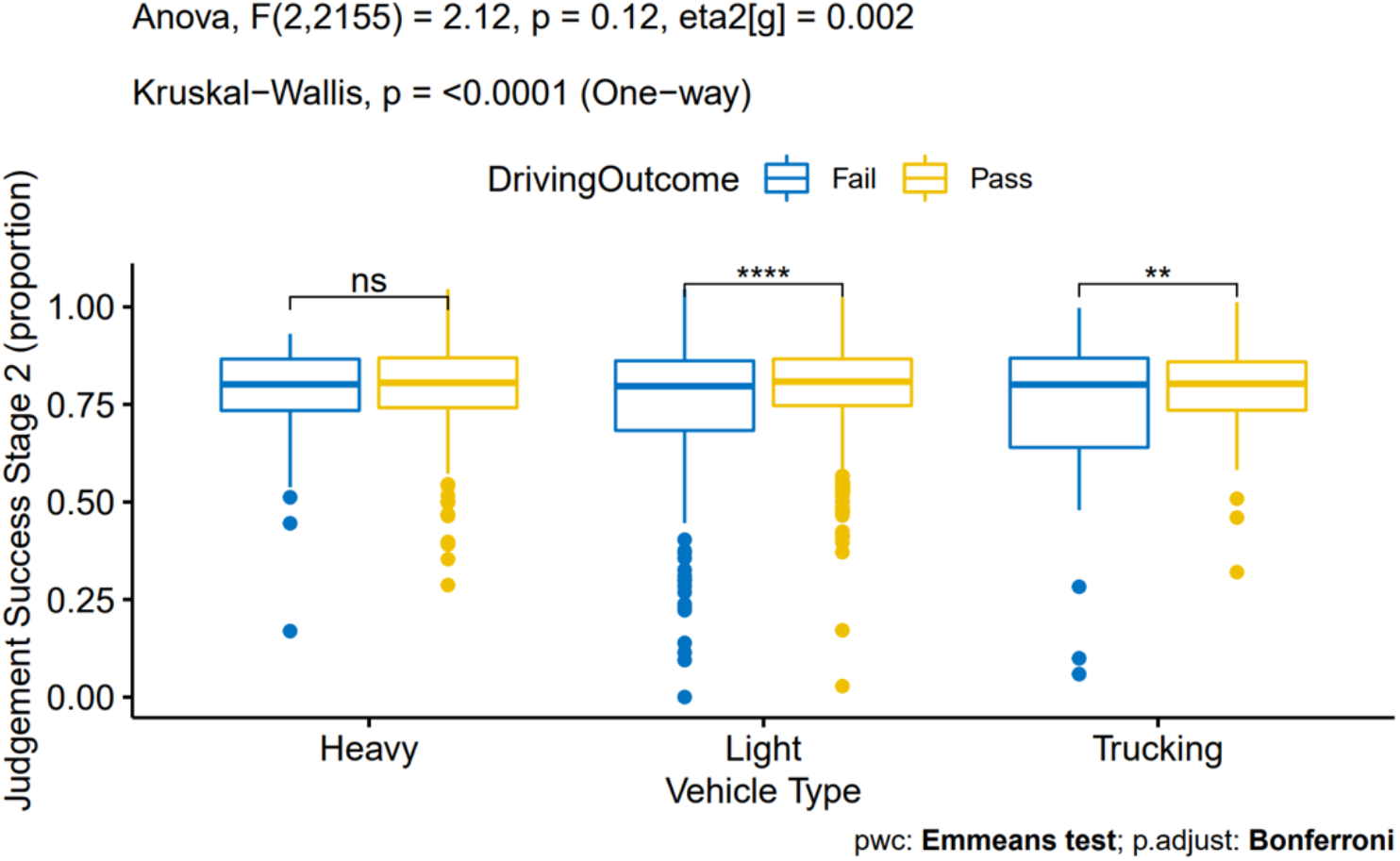
Group means and differences comparing success rate on Stage 2 of the Judgement task, depending on driving group and on-road test outcome. Light vehicles as well as truck drivers who passed the on-road test were significantly more successful than drivers who failed.

**Figure 7:**
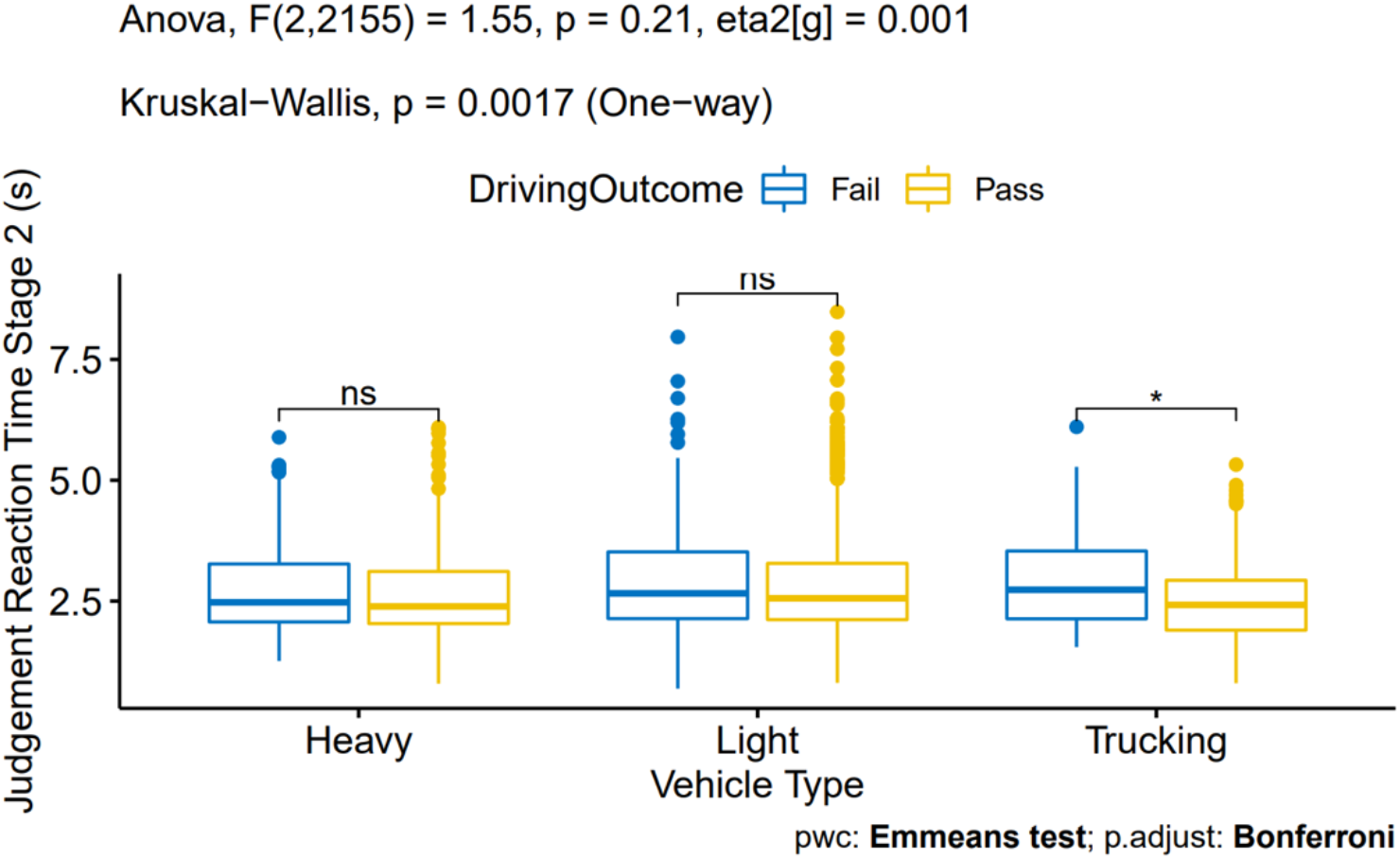
Group means and differences comparing reaction time on Stage 2 of the Judgement task, depending on driving group and on-road test outcome. Truck drivers who failed the on-road test were slower than truck drivers who passed.

**Figure 8:**
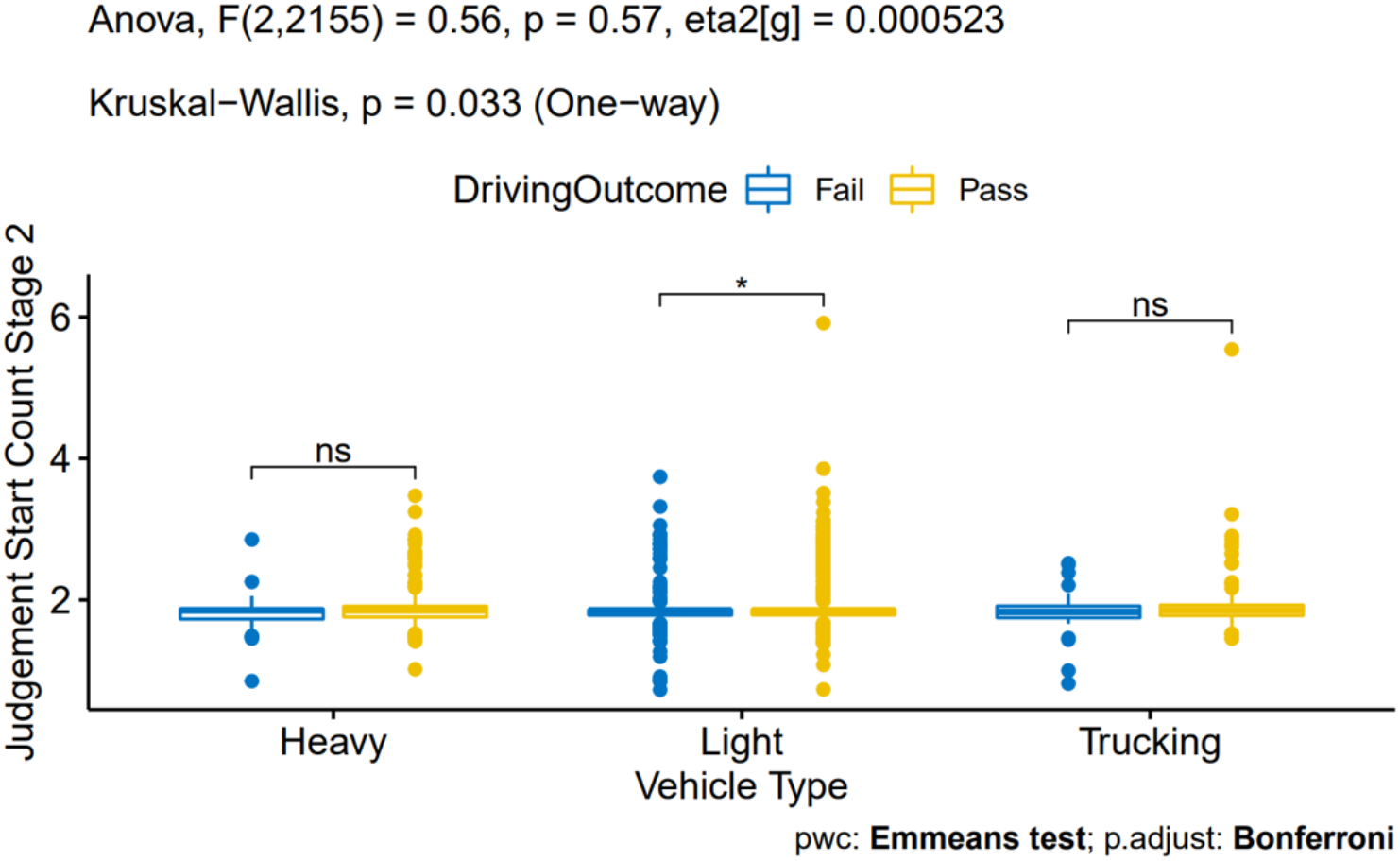
Group means and differences comparing the number of stops and starts to avoid obstacles on Stage 2 of the Judgement task, depending on driving group and on-road test outcome. Light vehicle drivers who passed the on-road test had more stops and starts than drivers who failed.

On the Memory task, light vehicle and truck drivers who passed the on-road test performed the task faster (Figure 9) and successfully replicated more shapes (Figure 10) than light vehicle and truck drivers who failed the test. No differences were observed for heavy vehicle drivers on either measure depending on if they passed or failed their test.

**Figure 9:**
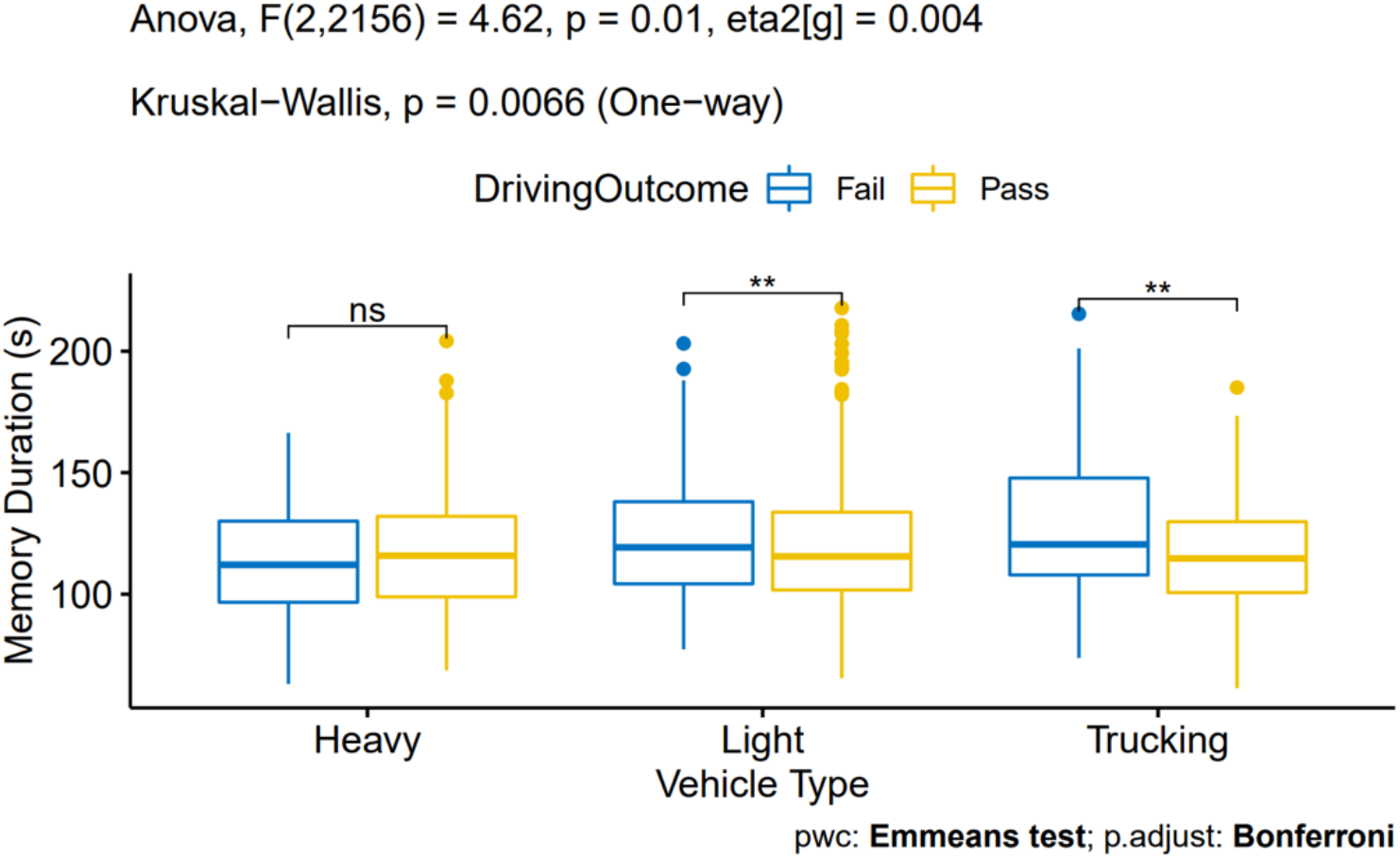
Group means and differences comparing trial duration on the Memory task, depending on driving group and on-road test outcome. Light vehicle drivers who failed the on-road test were slower to replicate the shapes than light vehicle drivers who passed. Truck drivers who passed the on-road test were faster to replicate the shapes than truck drivers who failed.

**Figure 10:**
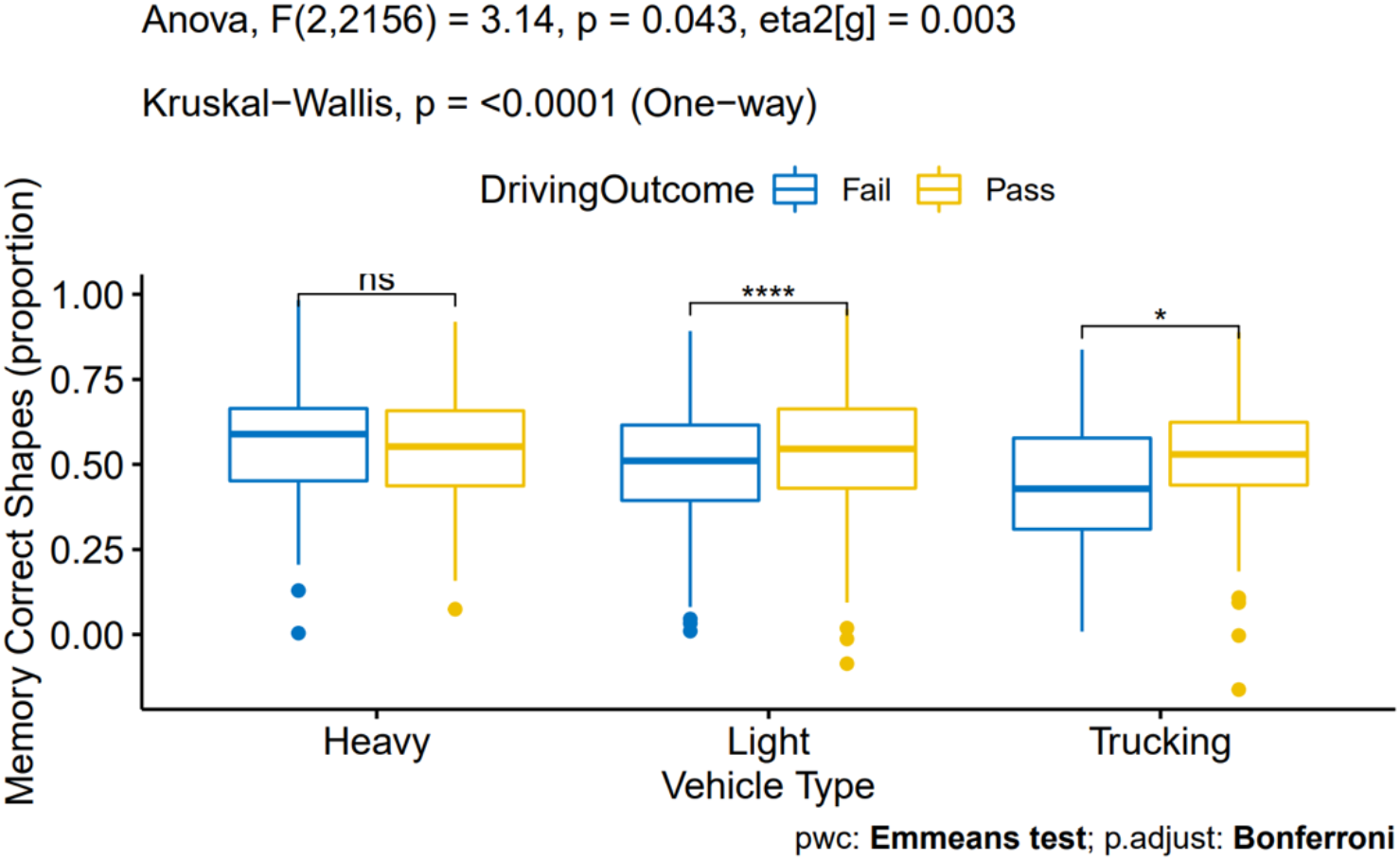
Group means and differences comparing proportion of total shapes that were successfully drawn on the Memory task, depending on driving group and on-road test outcome. Light vehicle drivers who passed the on-road test drew more successful shapes than light vehicle drivers who failed. Truck drivers who passed the on-road test drew more successful shapes than truck drivers who failed.

On the sensorimotor Control task, light and heavy vehicle drivers who passed the on-road test spent significantly more time within the target circle (Figure 11) compared to light and heavy vehicle drivers who failed, respectively. No differences were observed for truck drivers depending on if they passed or failed their test. No differences in the number of surprise obstacles hit were observed for any driving group, depending on if they passed or failed.

**Figure 11:**
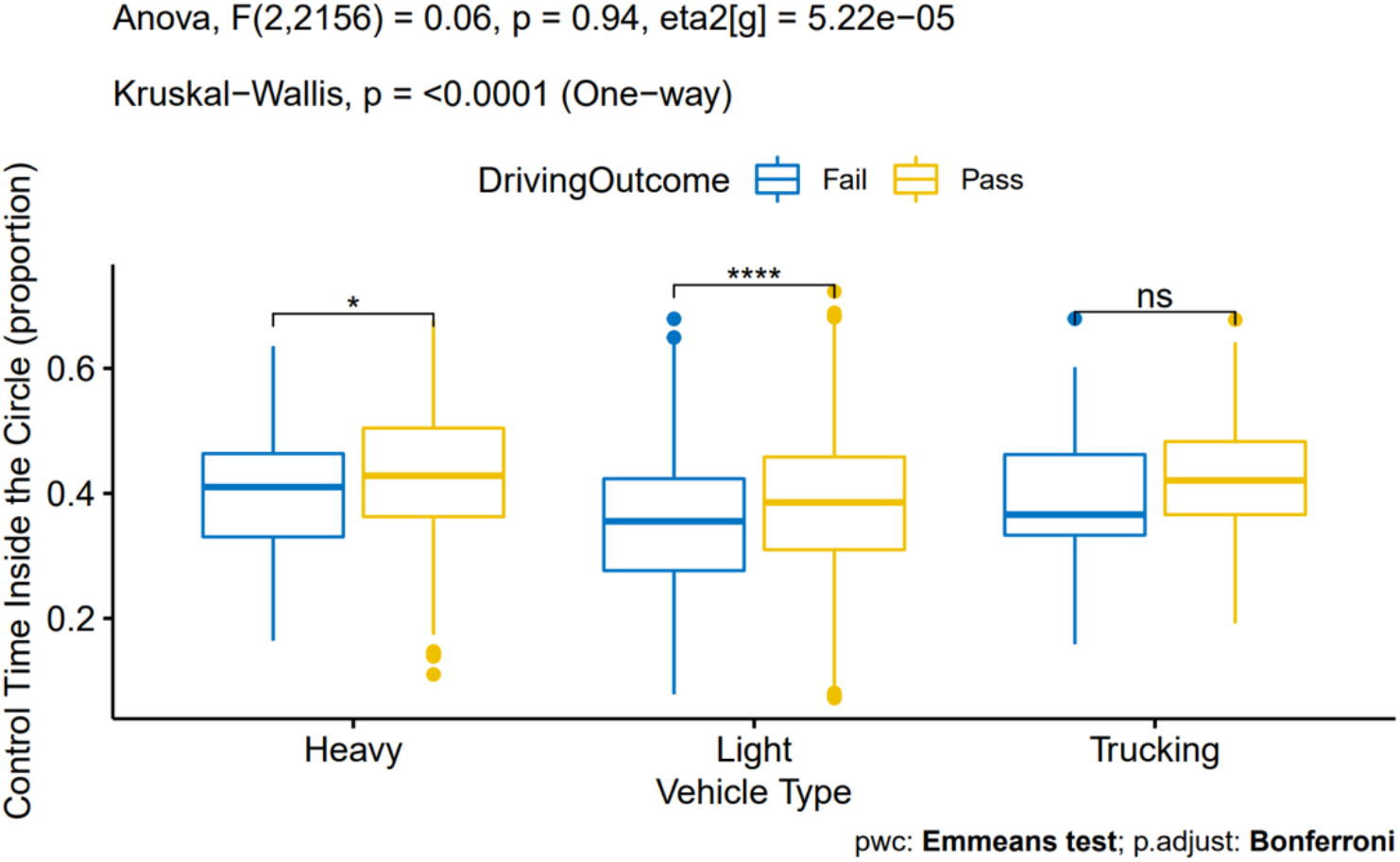
Group means and differences comparing proportion of total time on the sensorimotor Control task that was spent inside the target circle, depending on driving group and on-road test outcome. Light vehicles as well as bus drivers who passed the on-road test spent more time inside the circle than light vehicle and bus drivers who failed.

#### 3.2.1. Return-to-Work Group Excluded

Participants who were recruited into the study did so while completing the Vitals and an on-road driving test for a variety of employment-related reasons. A large majority of our participants were recruited as part of a pre-hire evaluation process, but some of our participants were already employed and were recruited into the study for other reasons, such as a routine wellness check, performance evaluation, or following an at-work safety incident (Figure 2). Some participants were also recruited as part of a return-to-work procedure and these participants were disproportionately represented in the Heavy vehicle group. In our study, there were 66 return-to-work participants in the Heavy vehicle group, compared to five and two such participants in the light and trucking groups, respectively (Figure 2d). Return-to-work participants were highly successful at passing their on-road test despite poor performance on the Vitals tool relative to other groups. To ensure that the over-representation of return-to-work participants in the Heavy vehicle group was not affecting the outcome of our group comparisons, we re-ran the univariate analyses from the previous section with the return-to-work participants excluded from the analysis. Table 6 shows the new sample sizes for each driving group with the return-to-work group excluded, along with the mean and median for each measure. Figures 12-20 show the ANOVA results for significant measures of the Vitals tool.

**Table 6:**
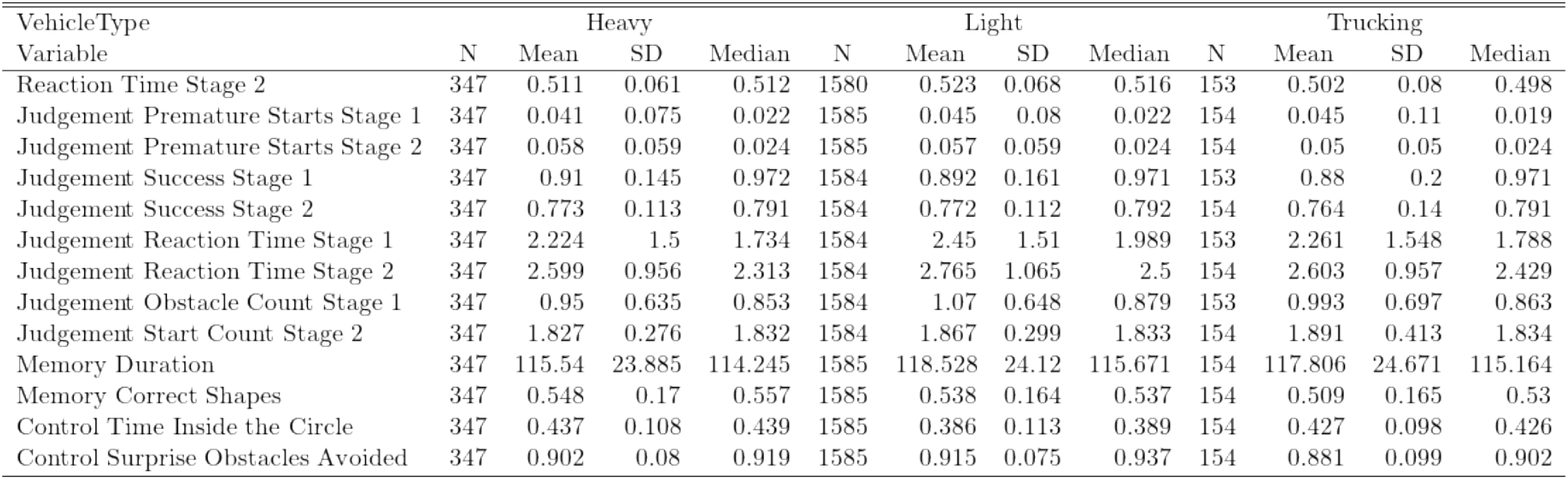
Mean and median performance on Vitals tasks by driving group (Heavy, Light, and Trucking) after excluding the return-to-work group (n=2091).

**Table 7:** Negative (NPV) and positive predictive value (PPV) of the classifier model validation dataset (n=2164).

| TN | TP | FN | FP | NPV | PPV | ACC | N (No Indeterminate) |
| --- | --- | --- | --- | --- | --- | --- | --- |
| 77.15% (1330) | 2.38% (41) | 17.0% (293) | 3.48% (60) | 0.8194 | 0.4059 | 0.7952 | 2164 (1724) |

**Table 8:** One-way ANOVA results on variables (with non-linear age effects removed) between high and low risk groups in the training dataset (n=1343).

| Variable | F | SS | $\eta_p^2$ | p |
| --- | --- | --- | --- | --- |
| Unnamed:0 | 17.9544 | 2.666935e+06 | 0.0132 | 0.0000 |
| Judgement1 | 40.8936 | 2.481698e+02 | 0.0296 | 0.0000 |
| Judgement2 | 26.8782 | 7.058030e+01 | 0.0196 | 0.0000 |
| Reaction3 | 8.6865 | 2.748900e+00 | 0.0064 | 0.0033 |
| Memory4 | 131.8788 | 4.061457e+02 | 0.0895 | 0.0000 |
| Memory5 | 107.7130 | 3.267430e+02 | 0.0744 | 0.0000 |
| Judgement6 | 47.9623 | 3.571164e+02 | 0.0345 | 0.0000 |

**Table 9:** Demographics of the training dataset (n=1343).

| | Age (M ± SD) | N | Gender (f/m) | Risk Level (Low/High) |
| --- | --- | --- | --- | --- |
| Low-Risk Group | 34.34 ± 17.23 | 955 | 397/557 | 0/955 (0.0%) |
| High-Risk Group | 42.66 ± 17.16 | 388 | 119/267 | 388/388 (100.0%) |
| Full Sample | 36.74 ± 17.61 | 1343 | 516/824 | 388/955 (28.9%) |
| Medical1 | 42.63 ± 15.53 | 93 | 19/74 | 3/93 (3.2%) |
| Research1 | 32.75 ± 5.18 | 99 | 47/52 | 0/99 (0.0%) |
| Research2 | 30.40 ± 9.43 | 269 | 129/140 | 123/269 (45.7%) |
| Medical2 | 74.21 ± 14.86 | 86 | 20/66 | 30/86 (34.9%) |
| Research3 | 20.31 ± 3.84 | 318 | 205/113 | 0/318 (0.0%) |
| Research4 | 44.97 ± 12.83 | 186 | 35/149 | 184/186 (98.9%) |
| Research5 | 69.64 ± 11.67 | 45 | 21/24 | 6/45 (13.3%) |
| Commercial1 | 39.98 ± 11.50 | 58 | 13/45 | 23/58 (39.7%) |
| Commercial2 | 38.31 ± 7.23 | 13 | 1/12 | 2/13 (15.4%) |
| Commercial3 | 38.65 ± 9.22 | 176 | 26/149 | 17/176 (9.7%) |

**Table 10:** Accuracy comparison of model output for various data sources in the training dataset (n=1343).

|  | ACC | FNR | FPR | TNR | TPR | Tri.Low | Tri.Up | Undefined |
| --- | --- | --- | --- | --- | --- | --- | --- | --- |
| Full-sample | 65% | 24% | 40% | 60% | 76% | 0.39 | 0.51 | 25% |
| Medical1 | 89% | 67% | 10% | 90% | 33% | 0.39 | 0.51 | 7% |
| Research1 | 84% | - | 16% | 84% | - | 0.39 | 0.51 | 36% |
| Research2 | 47% | 49% | 57% | 43% | 51% | 0.39 | 0.51 | 31% |
| Medical2 | 51% | 10% | 75% | 25% | 90% | 0.39 | 0.51 | 18% |
| Research3 | 69% | - | 31% | 69% | - | 0.39 | 0.51 | 26% |
| Research4 | 90% | 9% | 100% | 0% | 91% | 0.39 | 0.51 | 13% |
| Research5 | 26% | 20% | 82% | 18% | 80% | 0.39 | 0.51 | 18% |
| Commercial1 | 63% | 41% | 34% | 66% | 59% | 0.39 | 0.51 | 26% |
| Commercial2 | 71% | 50% | 20% | 80% | 50% | 0.39 | 0.51 | 86% |
| Commercial3 | 47% | 14% | 58% | 42% | 86% | 0.39 | 0.51 | 36% |

**Figure 12:**
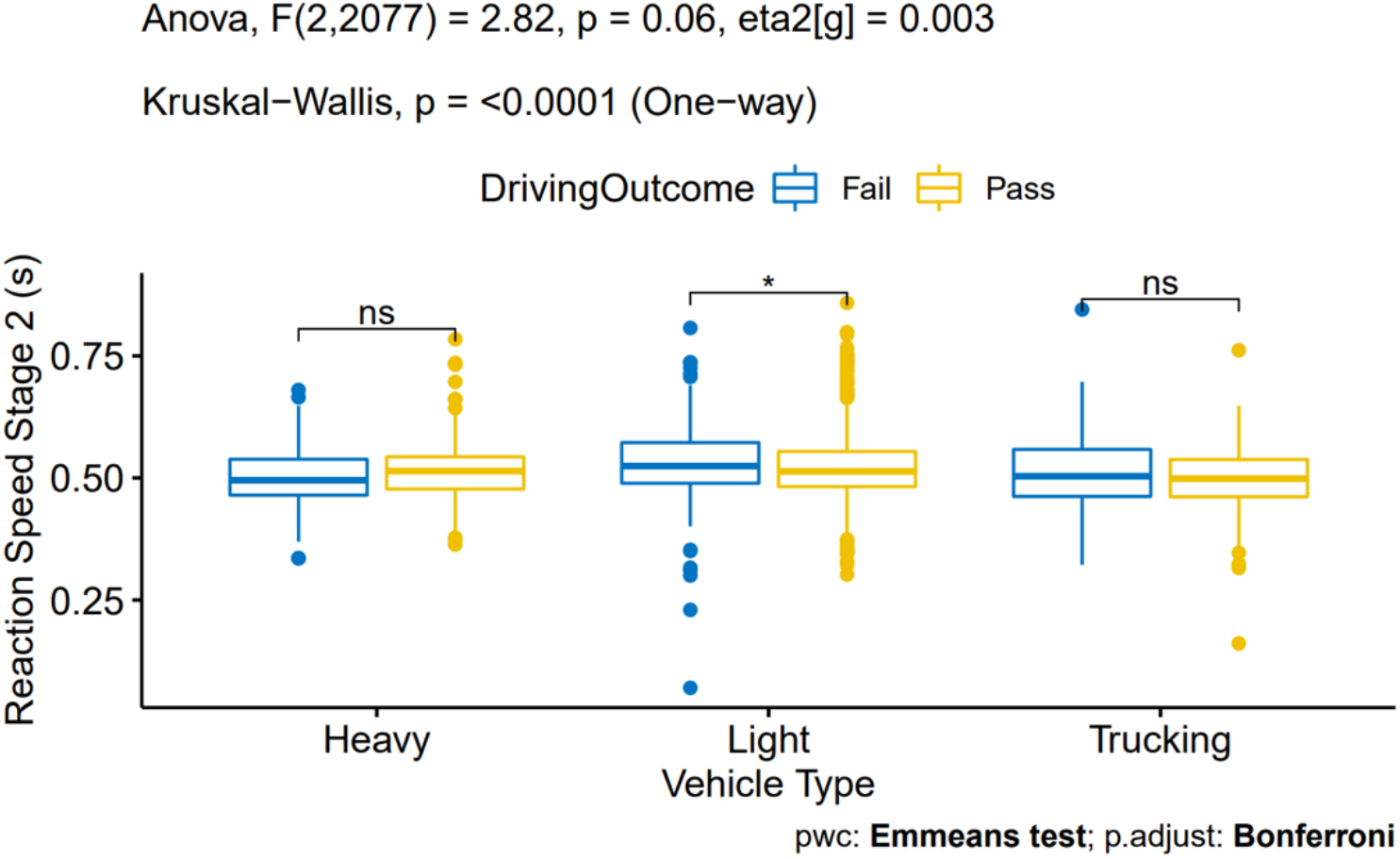
Group means and differences comparing reaction time on the RT task depending on driving group and on-road test outcome. Light vehicle drivers who failed the road test were slower than light vehicle drivers who passed.

**Figure 13:**
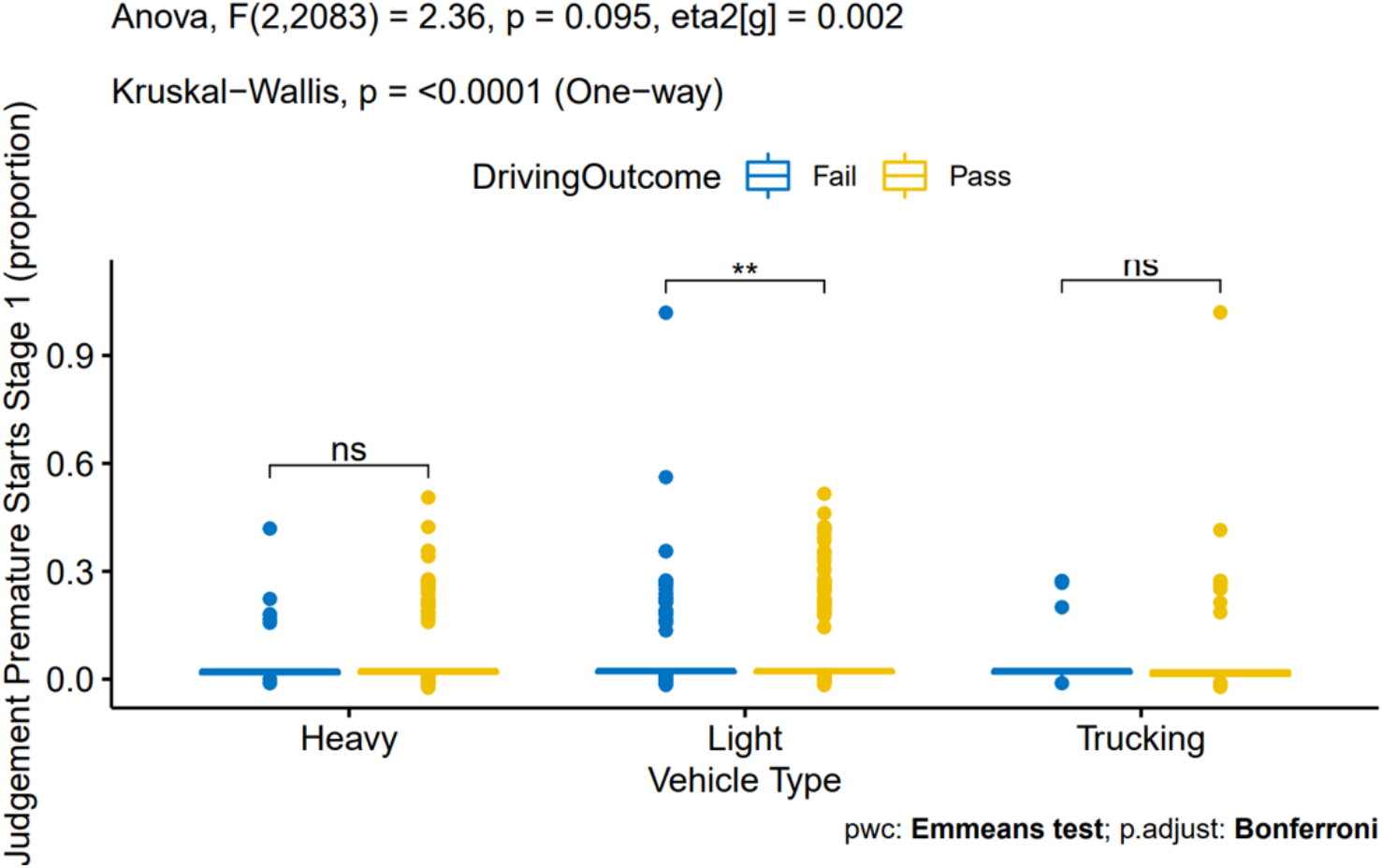
Group means and differences comparing proportion of total starts that were premature on Stage 1 of the Judgement task, depending on driving group and on-road test outcome. Light vehicle drivers who passed the test produced fewer false starts than light vehicle drivers who failed as a proportion of starts.

**Figure 14:**
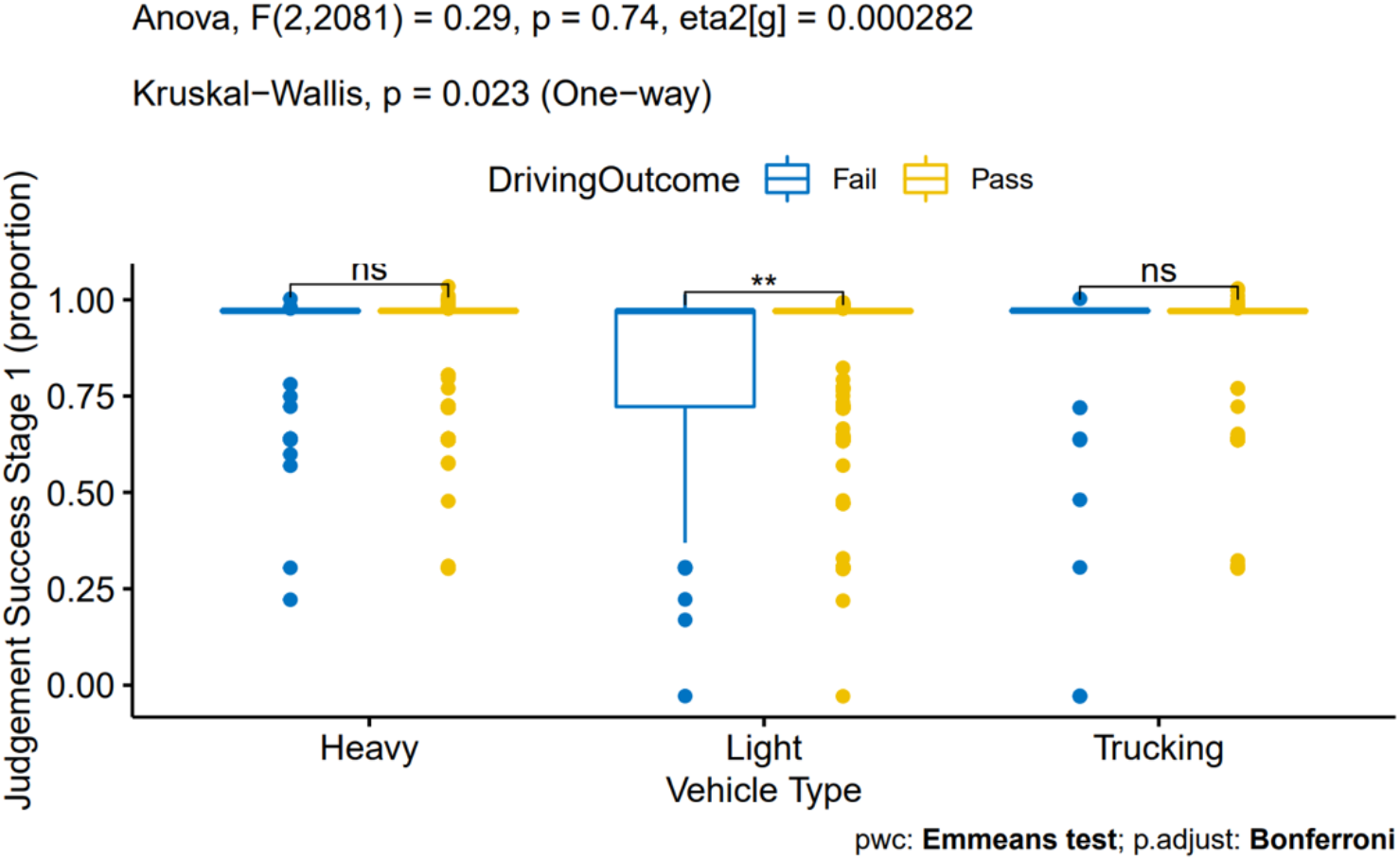
Group means and differences comparing trial success rate on Stage 1 of the Judgement task, depending on driving group and on-road test outcome. Light vehicle drivers who passed the on-road test were more successful than light vehicle drivers who failed.

**Figure 15:**
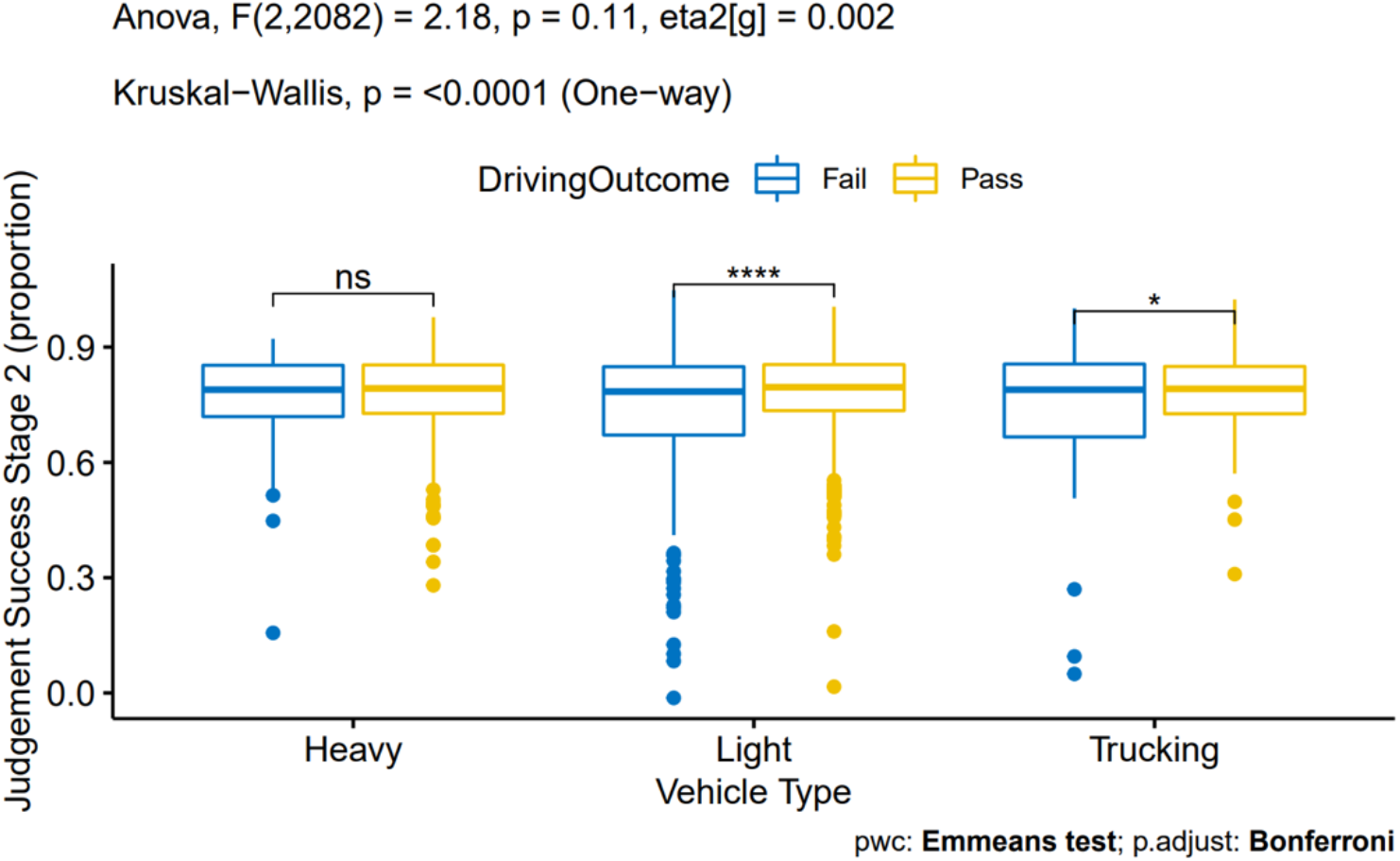
Group means and differences comparing success rate on Stage 2 of the Judgement task, depending on driving group and on-road test outcome. Light vehicles as well as truck drivers who passed the on-road test were significantly more successful than drivers who failed.

**Figure 16:**
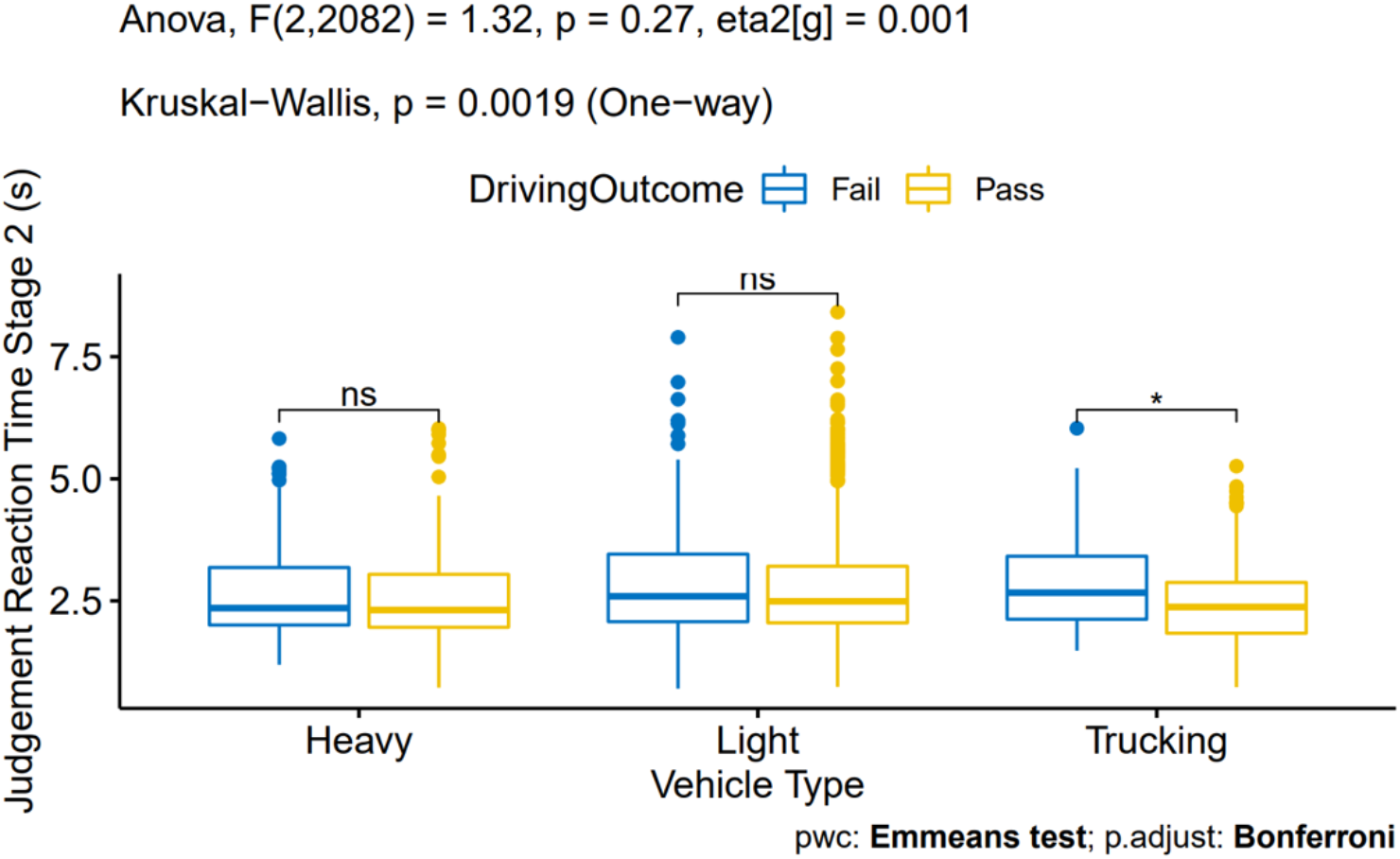
Group means and differences comparing reaction time on Stage 2 of the Judgement task, depending on driving group and on-road test outcome. Truck drivers who failed the on-road test were slower than truck drivers who passed.

**Figure 17:**
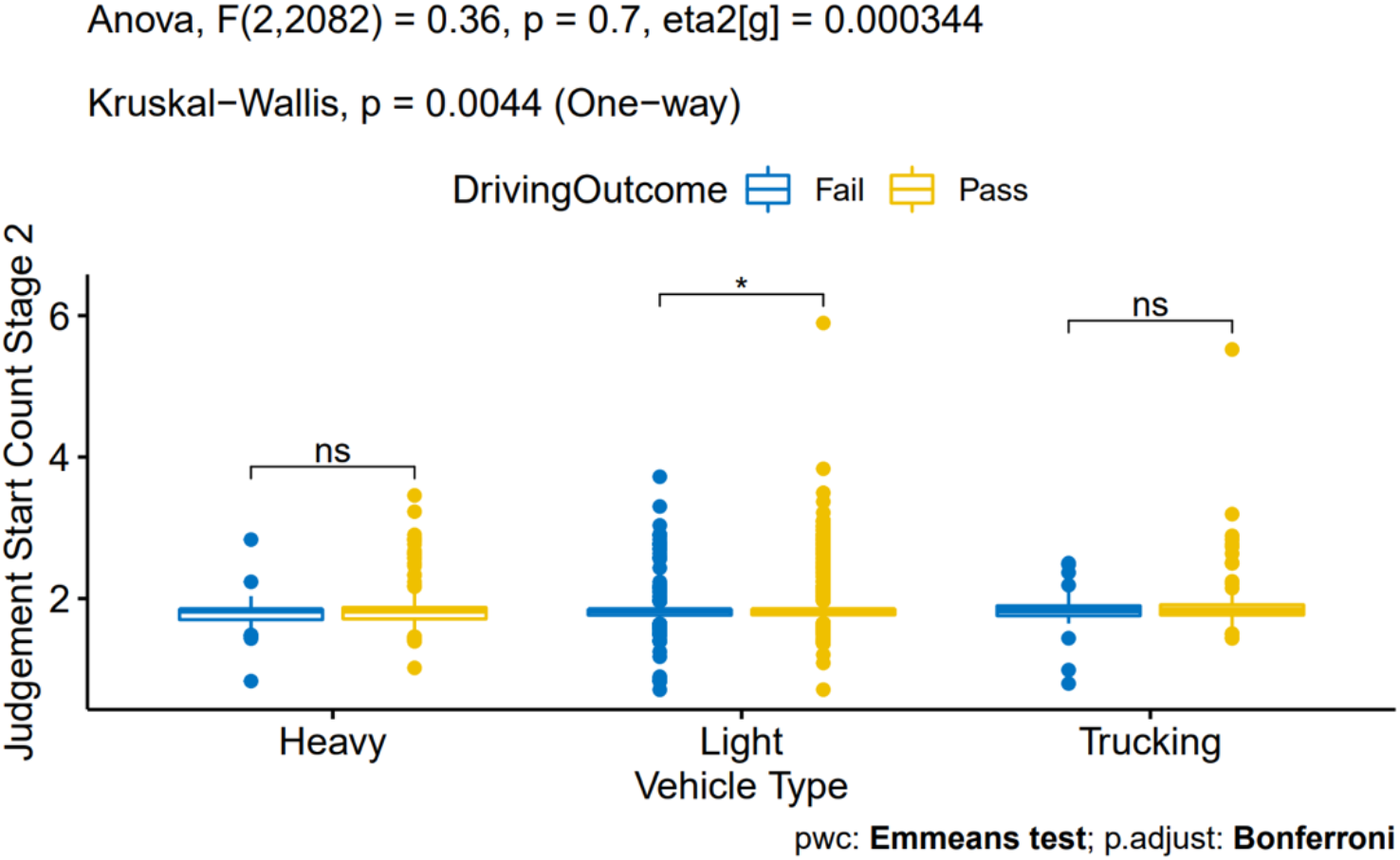
Group means and differences comparing the number of stops and starts to avoid obstacles on Stage 2 of the Judgement task, depending on driving group and on-road test outcome. Light vehicle drivers who passed the on-road test had more stops and starts than drivers who failed.

**Figure 18:**
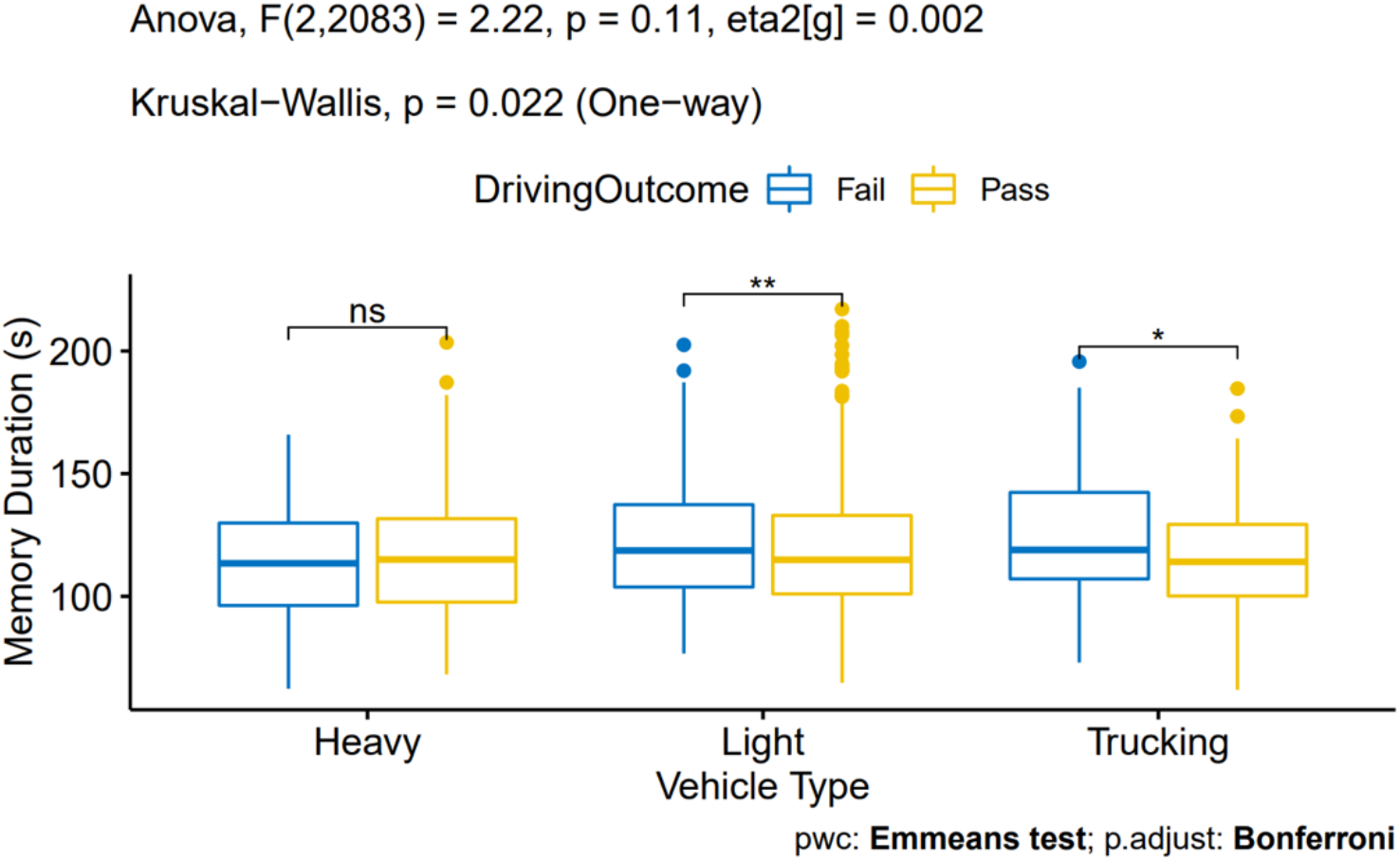
Group means and differences comparing trial duration on the Memory task, depending on driving group and on-road test outcome. Light vehicle drivers who failed the on-road test were slower to replicate the shape than light vehicle drivers who passed. Truck drivers who passed the on-road test were faster to replicate the shapes than truck drivers who failed.

**Figure 19:**
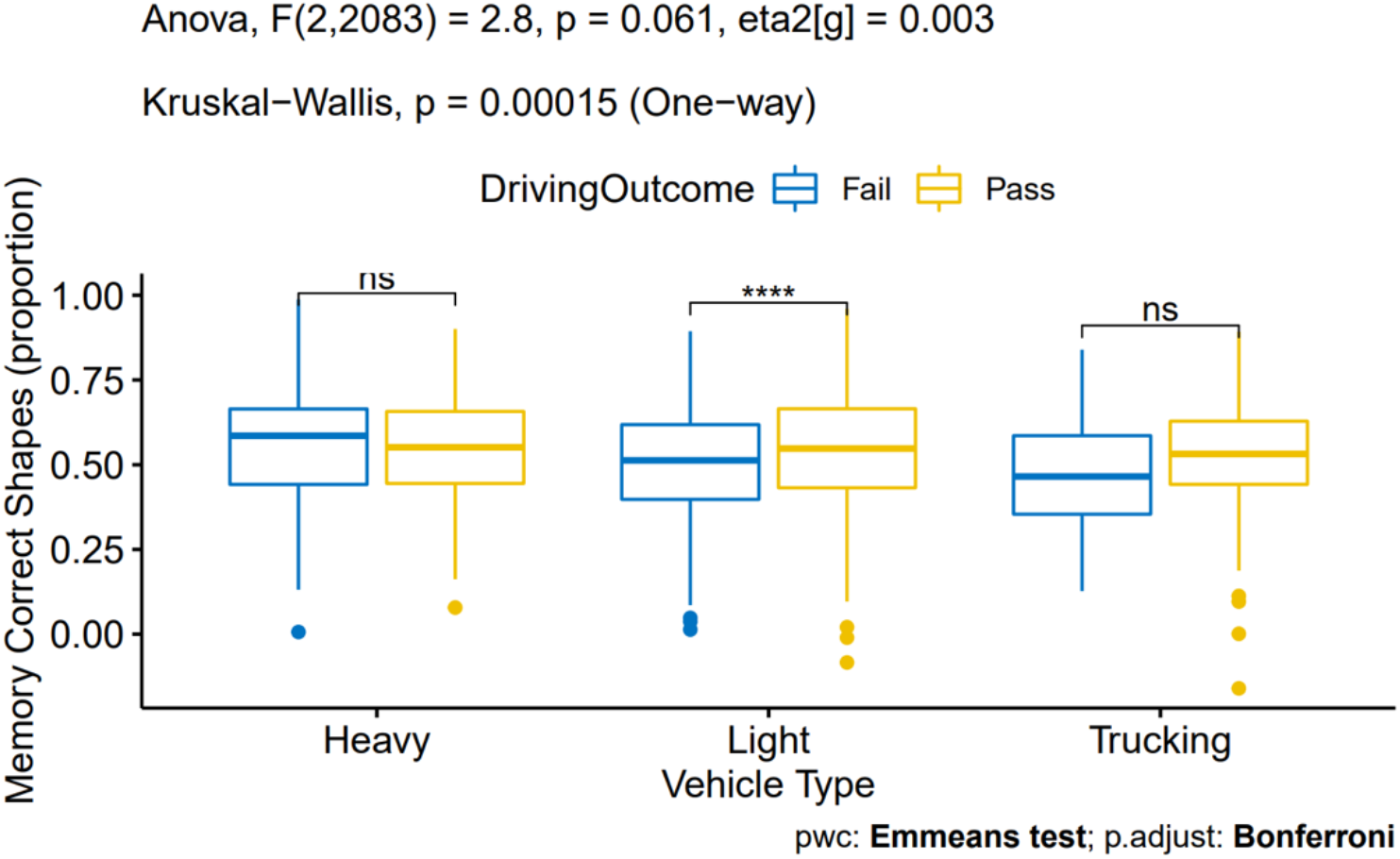
Group means and differences comparing proportion of total shapes that were successfully drawn on the Memory task, depending on driving group and on-road test outcome. Light vehicle drivers who passed the on-road test drew more successful shapes than light vehicle drivers who failed. Truck drivers who passed the on-road test drew more successful shapes than truck drivers who failed.

**Figure 20:**
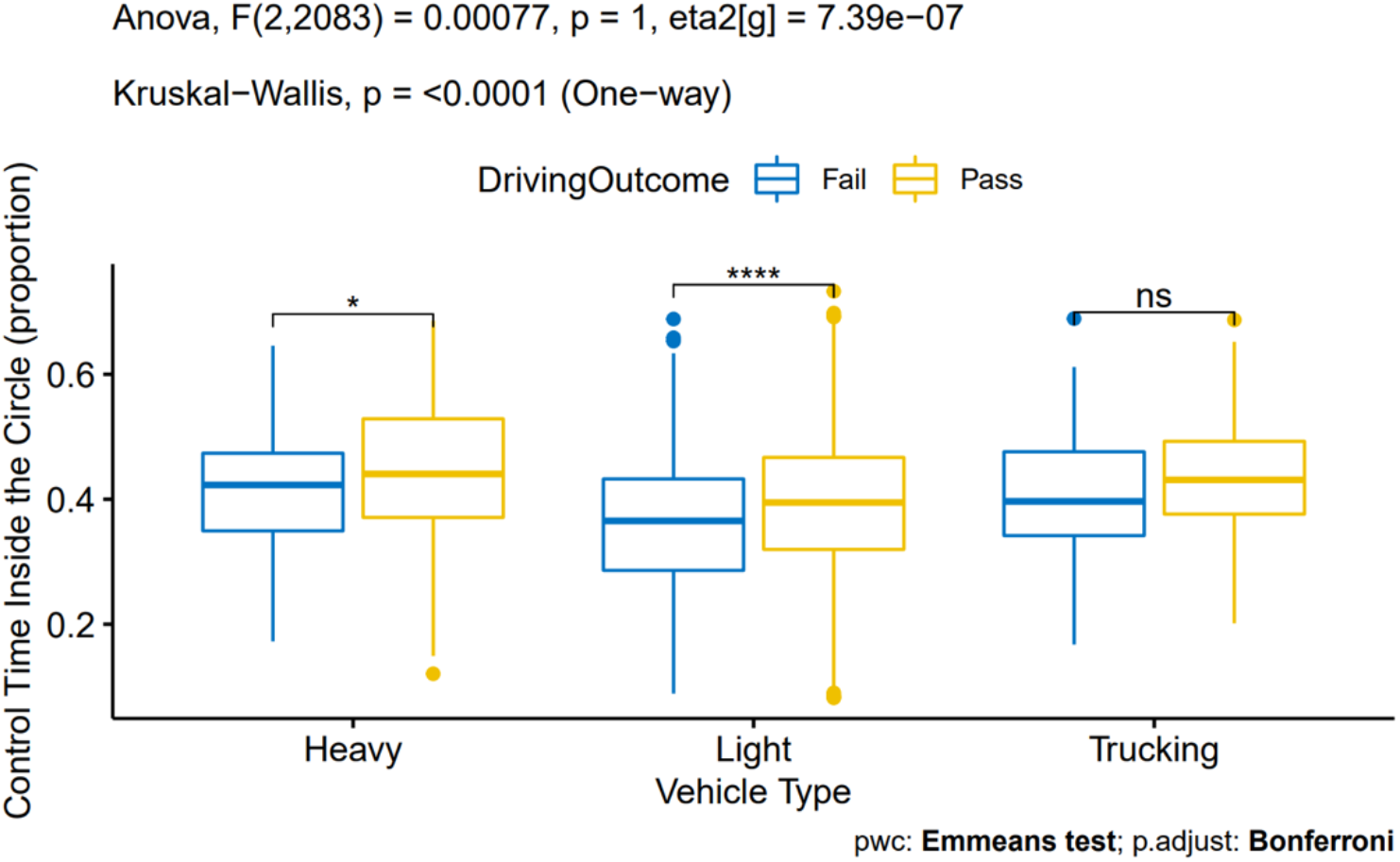
Group means and differences comparing proportion of total time on the sensorimotor Control task that was spent inside the target circle, depending on driving group and on-road test outcome. Light vehicles as well as bus drivers who passed the on-road test spent more time inside the circle than light vehicle and bus drivers who failed.

Excluding the return-to-work group from each ANOVA resulted in the outcome of several tests changing from statistically significant to non-significant, compared to the full sample. These tests included the RT task (Figure 12), the proportion of premature starts on the Judgement task in the second stage, trial duration on the Memory task (Figure 18), and proportion of successfully replicated shapes on the Memory task (Figure 19). For these four measures, excluding the return-to-work group meant that there were no longer any significant differences in Vitals performance depending upon driving group and on-road test outcome. Excluding the return-to-work group also resulted in a removal in statistical significance for one pairwise comparison, with the proportion of successfully replicated shapes on the Memory task no longer differing within the Trucking group depending upon their on-road test outcome (Figure 19).

### 3.3. Predictability of Vitals to Distinguish between Safe and Unsafe Drivers

Vitals data from this study’s participants was inputted into the model to predict whether participants would pass or fail their road test. Each participant in the study was assigned a score, calculated as the model’s predicted probability of their class being 1, i.e., “impaired”,. Likelihood of participants failing their on-road evaluation depending on their Vitals score is depicted in Figure 21. High-scoring participants were much more likely to fail the road test than low-scoring participants, showing the ability of the model to identify at-risk drivers on the basis of overall task performance.

**Figure 21:**
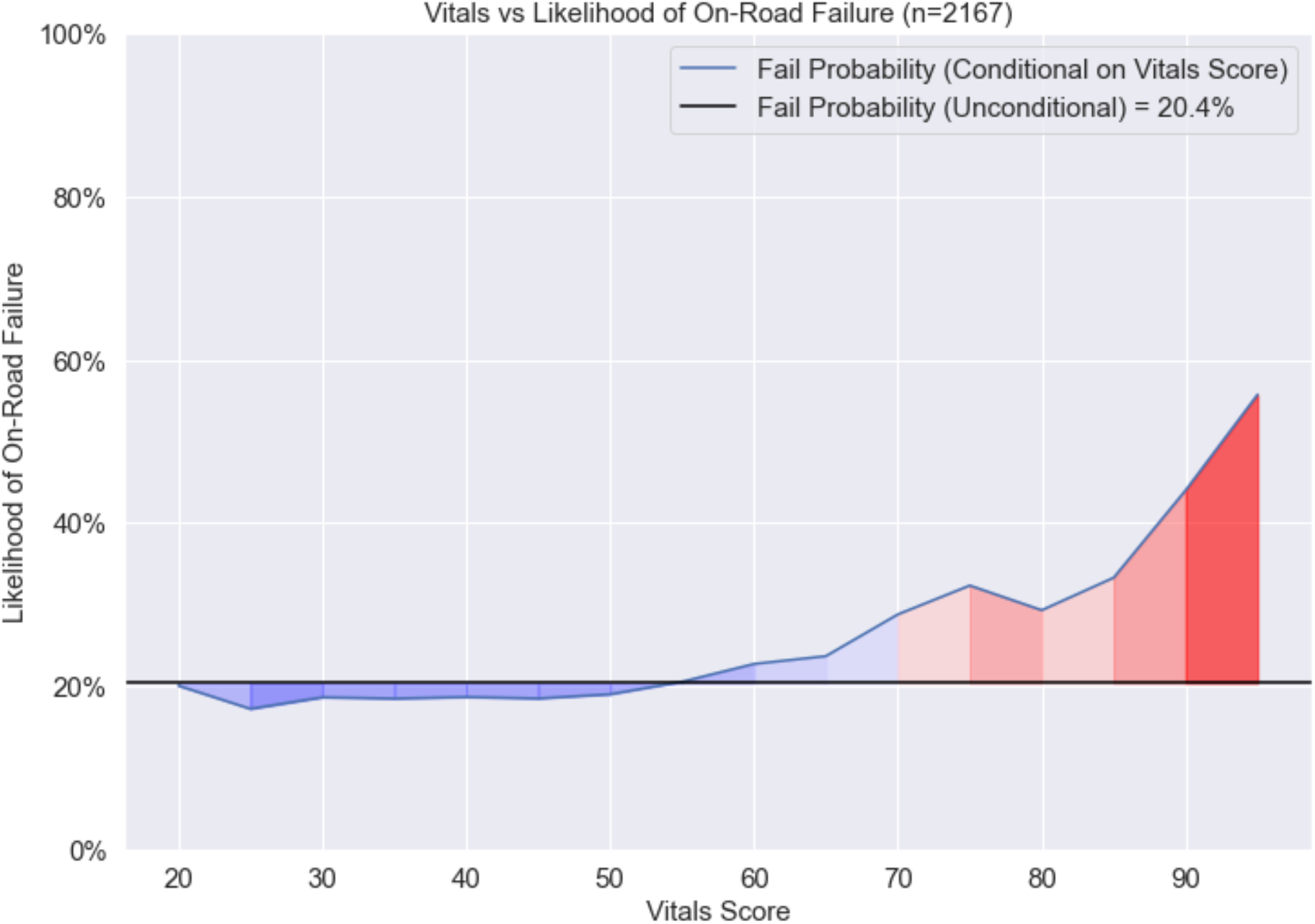
Likelihood of on-road test failure depending on Vitals score in the validation dataset (n=2167) using a smoothing width of 5 Vitals score points. Vitals score is calculated as the predicted probability of being impaired based on task performance,. A higher score indicates worse performance.

## 4. Discussion and Limitations

In the present study, we sought to examine performance on the Vitals task battery in a large sample of commercial drivers and compare across three vehicle types. The Vitals tool consists of four tests of reaction time, decision-making and judgement, memory, and sensorimotor control, and has been previously found to predict on-road driving performance in a group of healthy older adults (Dobbs et al., 1998), patients with dementia (Bakhtiari et al., 2020; Dobbs, 1997) or stroke (Choi et al., 2015) and in drivers under the influence of cannabis or cocaine (Tomczak et al., n.d.). In this study, we found that certain cognitive measures were better associated with passing an on-road commercial driving test, most notably simple RT, memory performance, and time-on-target in the Control task. This set of findings suggests that there is a strong relationship between these tablet-based tasks and real-world driving. Few differences were observed between the three driving groups, which may be because all three groups consisted of professionally trained drivers, and because truck and heavy vehicle drivers are highly likely to also be light vehicle drivers due to owning and operating a personal vehicle. It also stands to reason that the most sensitivity in the univariate analyses was associated with the light vehicle group since the sample size for this group was considerably higher than the other two groups.

### 4.1. Reaction Time

The Reaction Time task on the Vitals tool requires participants to observe two or three red lights flashing in sequence, followed by a flashing green light, and respond as quickly as possible once the green light is presented. This task conceptually resembles the experience of waiting for a traffic light to change before accelerating, and requires participants to engage in visual discrimination, sustained attention, response inhibition, and reaction time, among other neurocognitive abilities (Dobbs, 1997; Dobbs et al., 1998). Predictive modeling showed that faster reaction times were effective at specifying individuals who would go on to pass their on-road test. Furthermore, light vehicle drivers who passed their on-road test responded significantly faster compared to those who failed the test, according to an ANOVA analysis.

Reaction time is an extremely common measurement in cognitive and neuropsychological studies and is typically assumed to represent the speed (Pachella, 1973; Teichner, 1954) or stages of mental processing (Treisman, 1991). Slowed reaction times are often seen in individuals who have experienced a head injury (Balaban et al. 2016; Stuss et al., 1989), are physically frail (Robertson et al., 2014), or are experiencing pathological aging processes, such as mild cognitive impairment and Alzheimer’s Disease (Gorus et al., 2008). It is also known that reaction time performance generally peaks in early adulthood before declining and becoming more variable as people age (Hultsch et al., 2002; Tun & Lachman, 2008). Reaction time is a very important aspect of safe and successful driving, as drivers are often required to accelerate or decelerate their vehicles in response to visual stimuli, such as traffic lights, obstructions and debris, or the behaviour of other road users. Reaction time is especially important for decelerating a moving vehicle, as a slower reaction time will translate into longer stopping distances and greater momentum at impact, with potentially serious or even fatal consequences. Given this, it should be unsurprising for a test of reaction time to demonstrate value in predicting whether a driver will pass or fail a road test. Drivers who exhibit slow reaction times are more likely to respond slowly or improperly to road signals and to other road users, with a corresponding increase in safety risk to themselves and others. It is also the case that slower reaction times are generally associated with more response variability from trial-to-trial, which may be related to overall poorer on-road performance compared to drivers with less variability in simple motor responses, like braking.

### 4.2. Decision-Making

The Judgement task requires participants to control a virtual icon and safely cross it from one side of the screen to the other using a Start/Stop button without contacting any obstacles. The task consisted of two levels of difficulty: In Stage 1, participants had to navigate across one lane of “traffic flow,” which consisted of rising or falling blocks separated by navigable gaps, while in Stage 2, participants had to navigate across two lanes of flow moving in opposite direction to one another. This task conceptually resembles the act of navigating a vehicle across one or two lanes of traffic and collectively measures several executive functions such as attention, decision-making, and reaction time, as well as sensorimotor abilities such as judging and predicting the motion of objects (Dobbs, 1997; Dobbs et al., 1998).

Contrary to the RT task, reaction time in the Judgement task was not associated with on-road test outcome. The Judgement task is more complex than the RT task, and participants are responding to and attempting to navigate a flow of sequential stimuli, rather than reacting to a single stimulus. Depending on the location of stimuli at trial onset, a faster reaction time may in fact be contrary to successful task performance and lead to a collision, whereas waiting for a gap in the flow of stimuli to make a response is appropriate. Thus, reaction time is not as good a proxy for driving ability on this task as other measures may be. Moreover, the Judgment task requires higher-order cognition associated with making and implementing a decision about moving obstacle avoidance. Such a task can involve different strategies, where one driver may choose to take a more cautious approach even if it does not necessarily lead to a safer outcome. Thus, the different strategies could be associated with different response times but not different outcomes, which could work against this measure being predictive of on-road success.

### 4.3. Short-Term Memory

To complete the Memory task component of the Vitals tool, participants are shown a geometric shape and then asked to recreate it after a brief delay. According to our classifier model, task duration and replication success were both good predictors of future on-road test outcomes. Participants who correctly replicated more shapes and took less time to complete the task were more likely to pass their road test than participants who replicated fewer shapes and took longer to complete the task. Between-group comparisons showed significant differences on both measures for light vehicle and truck drivers but not heavy vehicle drivers. The differences in statistical significance between the light vehicle group and the other groups are likely due to the much larger sample size for the light vehicle group, meaning that they better reflect the true population means for the pass and fail groups.

Overall, memory is a well-established proxy for general cognitive functioning. Working memory involves the ability to flexibly coordinate other mental abilities, be they semantic, sensory, or motoric (Baddeley, 1992; D’Esposito & Postle, 2015), and therefore poor or declining working memory function can have broad impacts on an individual’s ability to perform complex tasks. Memory impairments are a common symptom of mild cognitive impairment as well as neurovascular or metabolic conditions such as stroke or diabetes, and declining memory function is often the first observable symptom of neurodegenerative conditions such as Alzheimer’s disease (Alzheimer’s Association, 2022). In the case of MCI and dementia, symptoms primarily affect episodic or short-term working memory rather than semantic or long-term memory. Accordingly, tests of short-term or working memory, such as the Mini-Mental State Exam and the digit span task, are often employed by physicians for diagnostic purposes (Creavin et al., 2016; Huntley & Howard, 2010). Our results indicate that performance on a short-term memory task is predictive of driving ability and likelihood of impairment. Driving involves regular use of short-term, working memory functions, such as maintaining in one’s mind the information posted on road signs, or the locations of other road users. Consequently, a driver whose short-term, working memory performance is measured to be low or declining may be a strong candidate to recommend for an on-road test of their driving ability. This result may also have some diagnostic utility, as it suggests that people who score low on the Vitals Memory task may be sufficiently impaired for this to interfere with daily functioning, including but potentially not limited to, driving.

### 4.4. Sensorimotor Control

The final component of the Vitals tool, the sensorimotor Control task, requires participants to maneuver a virtual rolling ball inside a target circle by tilting the tablet left or right like a steering wheel, while avoiding any sudden obstacles that appear. According to our model, time spent inside the target circle predicted future on-road test outcomes, but obstacle avoidance did not. The Control task is a complex task which, like driving itself, requires a wide variety of perceptual, cognitive, and motor skills in order to perform skillfully (CCMTA, 2022). In this task, participants must use visual pursuit to follow the target as it moves across the screen, perform fluid movements to maintain the ball within the moving target, and automatically orient towards and react quickly to stimuli, among other skills. Decision-making during the task can either be good-based, such as whether to stay in the circle or turn to avoid an obstacle depending upon the relative value placed on each outcome, or action-based, such as whether to turn left or right to avoid a sudden obstacle depending upon the relative spatial locations of the ball, target, and obstacle (Wispinski et al., 2020). Given the complexity of the task, there may be no clear and simple explanation for why time in the circle but not obstacle avoidance predicts on-road test outcomes. Nevertheless, we can still offer some speculation and lay the groundwork for future study.

This pattern of results further supports the relationship between sensorimotor processing and driving. It is not surprising that the time-on-target in this task varied as a function of pass/fail on the driving test. The Control task relies on a series of cognitive processes and, importantly, also relies on motor control in a similar manner to steering wheel operation. Using fine motor movements to maintain one’s vehicle within a visual lane is a critical driving skill that is continuously required of drivers. By contrast, sudden obstacle avoidance is still critical but less frequently required and is not deliberately tested during an on-road evaluation for obvious safety reasons. Failing to avoid an obstacle is also much less critical during the Control task than it would be while operating a vehicle, and participants may have simply chosen to prioritize increased time on target over successful obstacle avoidance, leading to a dissociation between motor control skill and performance on this measure. Therefore, performance at avoiding obstacles on the Control task may not relate to performance during a road test, despite being theoretically relevant to real-world safe driving.

### 4.5. Limitations

A real-world study of this nature is not without challenges and limitations. Data sources for the classifier model had unequal sample sizes, and oversampling of small sources could not be used since some sample sizes were very small. Thus, all sources were weighted equally when building the model. Participant demographic data for the study did not include information on gender. Precise vehicle models operated by each participant were not collected (participants were sorted into groups based on the source of their data), so each vehicle category (Light, Heavy, and Trucking) contains various vehicle models which may differ in their utility and physical dimensions. Participants in the heavy vehicle group were disproportionately recruited to the study via return-to-work procedures after a safety incident at work.

## 5. Conclusions

In this study, we aimed to predict on-road driver performance in commercial heavy vehicle and truck drivers, as well as light vehicle drivers, depending on performance on the tablet-based Vitals cognitive screening tool. On-road driving evaluations are expensive, time-consuming, and can be unsafe for drivers who are most at-risk, meaning that a reliable off-road screening tool can have substantial benefits for both drivers as well as the organizations or jurisdictions that administer the evaluation. The results of the study showed that performance on the Vitals was effective in predicting on-road performance across all three groups. Furthermore, drivers who failed their on-road evaluation performed significantly worse on several measures of the Vitals, including Judgement task success, correct shape replication on the Memory task, and time on-target in the Control task, compared to drivers who passed. This is the first study to demonstrate the Vitals’ effectiveness among commercially licensed drivers. Whether drivers are seeking to renew their license or resume driving after a health or safety incident, the use of the Vitals as a screening tool can benefit both commercial vehicle drivers and evaluators in distinguishing between drivers who are likely to pass or fail an on-road evaluation.

## Data Availability

All data produced in the present study are the confidential property of Impirica.

